# Prognostic Features of Anti-Cancer Drugs Response in Resected/Unresected Primary Non-Small Cell Lung Cancer: A Retrospective Cohort Study

**DOI:** 10.64898/2026.07.07.26357288

**Authors:** Saikat Samadder

## Abstract

**Aim:** Low chemotherapy response is a major risk factor for early mortality in cancer patients; it is one of the biggest challenges in cancer treatment. Main aim of this study is to identify chemotherapy non-responder, prognostic significance of pre-chemotherapy baseline variables in survival, distinguish most effective anti-cancer drug classes and formulation.

**Methods:** In this multi-center retrospective cohort (n=2459) patients deceased with NSCLC and received anti-cancer drugs were included for analyses. To identify chemotherapy non-responder, patient population was divided into three sub-groups based on chemotherapy prescription frequency [1–15] as group-A, [16–30] as group-B, and [≥31] as group-C. Multivariate analysis was performed to identify risk of 1-year mortality in these groups. To prognose chemotherapy response in resected and unresected NSCLC patients, 0-7 days pre-chemotherapy white blood cell (WBC) count total five-ranges were compared as per overall survival in abnormal Vs normal WBC counts.

**Results:** Post-stratification in group-A there were (n=1289) patients, in group-B (n=648) patients, and in group-C (n=522) patients. In group-A (n=301) patients 23% were found to have no new metastasis post-diagnosis significantly less *p*-value (0.004) compared to Group-B (n=125) 19.3%, and group-C (n=110) 19.2% patients *p*-value (0.008). Metastasis during chemotherapy was found significantly less in 20% patients of group-A, compared to (33%) in group-B, and (43%) in group-C *p*-value (<0.001). Post-chemotherapy initiation OS in group-A patients were significantly less 9 months (95% CI 9.3 – 9.6) compared to group-B 19 months (95% CI 17.7 – 20.2) and group-C 36.6 months (95% CI 34.6 – 38.5) patients p-value (<0.0001). Despite of low new metastasis and post chemo metastasis, group-A patients survived significantly less based on these outcomes group-A patients were considered as chemotherapy non-responder. Males and NSCLC stage III/IV patients were at higher risk; clinical benefits are corelated to surgery and radiotherapy for chemotherapy non-responder. Leukocytosis in both resected/unresected NSCLC group-A (13%) patients were found to be bad prognostic factor of survival in unresected group-B (5%) patients. Oral formulation of receptor tyrosine kinase inhibitors (RTKI) was effective in non-responders.

**Conclusion:** Stratification of patient population based on chemotherapy prescriptions could be a useful method to find chemotherapy response in retrospective analysis. Patients with pre-chemotherapy leukocytosis should be closely monitored prior to selection of chemotherapy dose and formulation.

## Introduction

Lung cancer is one of the main cause of deaths in global cancer population, majority of lung cancer patients (85%) are diagnosed with non-small cell lung cancer [1–2]. There are several classes of anti-cancer drugs including chemotherapy, immunotherapy, kinase inhibitors, and targeted therapy available for treatment of NSCLC patients [3]. Adjuvant chemotherapy is more beneficial for NSCLC compared to neoadjuvant chemotherapy, although outcome largely depends on diagnosis at early stage [4–6]. First-line chemotherapy combination of cisplatin/carboplatin with paclitaxel were reported as best first-line for NSCLC treatment compared to combination of cisplatin with vinorelbine [7–9]. Adjuvant chemotherapy selection during initial screening was based on gender, age, cancer stage, tumor grade, body weight, height, and performance status reported in an observational study [10].

Anti-cancer drug resistance is one of the biggest challenges in cancer treatment, despite of increased biomarkers, and drug targets discovered in past decades [11–12]. A recent article reported that more than 90% (±5%) cancer patients failed anti-cancer drugs, suggested by industry and regulatory experts [3, 12]. Toxicities contributed by anti-cancer drugs are the main reason of treatment discontinuation causing rapid cancer progression in patients [8]. There are several drugs available to minimize low or medium grade toxicities allowing to continue chemotherapeutic regimen safely [13–15]. However, in end-stage of cancer hematological toxicities, infections, organ failures are the major cause of fatalities contributed by extensive or intensive regimen of chemotherapy, irrespective of age as contributing factor [8]. Mortality in several cancer patients could be shortly after chemotherapy initiation or within a month after low dose chemotherapy initiation irrespective of chemotherapy dose or combinations [16]. These patients are non-responders of first-line chemotherapy, are under constant threat of severe toxicities resulting in early mortalities.

Chemotherapy non-responders are poorly reported across medical literatures mainly in retrospective studies, very few studies are currently available [16–18]. Tumor size increase or decrease as outcome measure of clinical trials may not be corelated with overall survival were demonstrated earlier [19–20]. Gastric cancer patients with at-least (-)30% decrease in tumor size were found to be complete responder of chemotherapeutic agents showed increased overall survival in clinical trials [21]. Multivariate analysis of variables such as surgery, metastatic sites, metastasis during chemotherapy had no statistical significance among progressive disease plus stable disease patients as combined group compared to partial response group, clinical trial drug response were mainly based on RESIST (1.0 or 1.1) [21–22]. Whereas cancer stage and tumor grade at baseline are often correlated with hazard ratio in retrospective studies and mainly reported to prognose survival outcome [23]. This may not be directly related to longitudinal chemotherapy response since at the end of each line of chemotherapy mortalities are prominent due to poor chemotherapy response, previously shown related to baseline ECOG ≥2 and smoking status in unresected NSCLC patients [24]. Earlier age and comorbidity were found to be independent predictor of treatment response in NSCLC patients [25]. Apart from tumor response, cancer drugs are approved based on median overall survival (OS) and progression-free survival (PFS), mean OS/PFS was suggested for regulatory reporting purpose to avoid over estimation [26].

Prognosis of chemotherapy response was previously studied in animal models using ultrasound as guiding tool for prognosis of chemotherapy response [27], (18)F-fluorodeoxyglucose ((18)F-FDG) reuptake in PET/CT was considered to be prognostic factor for chemotherapy response [28–29]. Glasglow prognostic score at baseline used abnormal CRP of ⩾10mg/L and hypoalbuminemia of <35mg/L were not useful for prognosis of chemotherapy response in NSCLC patients [30]. While baseline hematological markers lymphocytes, WBC, polymorphonuclear leukocyte, and platelet counts were referred in humans for prognosis of post-chemotherapy early mortalities [18]. WBC count of above 7x10^3^ cells per ml of blood obtained during first-line adjuvant chemotherapy was found to be bad prognostic factor for overall survival in NSCLC patients [31–32]. The main purpose of this rare multicenter retrospective cohort study is to identify patients with low/poor chemotherapy response, associated risks of 1-year mortality, and prognosis based on survival outcome as per WBC counts collected 7 days pre-chemotherapy. This study compared post chemotherapy survival outcome in responders and non-responders treated with various types of anti-cancer drug classes and formulation types such as oral, injections and oral plus injections. This study highlighted few important clinical practice to improve chemotherapy response in non-responders.

## Materials and methods

### Study Design

In this multi-center retrospective cohort study electronic medical records (EMR) of three branches of Severance hospital, Seoul were analyzed. There are (n=7532) patients diagnosed with lung cancer from 1^st^ Jan 2010 to 1^st^ Jan 2020 was assessed to identify primary lung cancer patients treated with anti-cancer agents. Lung cancer patients (n= 3985) excluded based on lost to follow-up or not deceased until 30^th^ June 2024, patients with secondary lung cancer diagnosed at the time of first cancer screening (n=750), diagnosed with small-cell lung cancer patients (n=335) or (n=3) mesothelioma. Application of this exclusion criteria, identified (n=2459) deceased patients as primary non-small cell lung cancer were included for further analyses to generate real-world evidence from EMR data. Flowchart for NSCLC patient selection and stratification is available in **Figure 1**.

**Figure 1:**
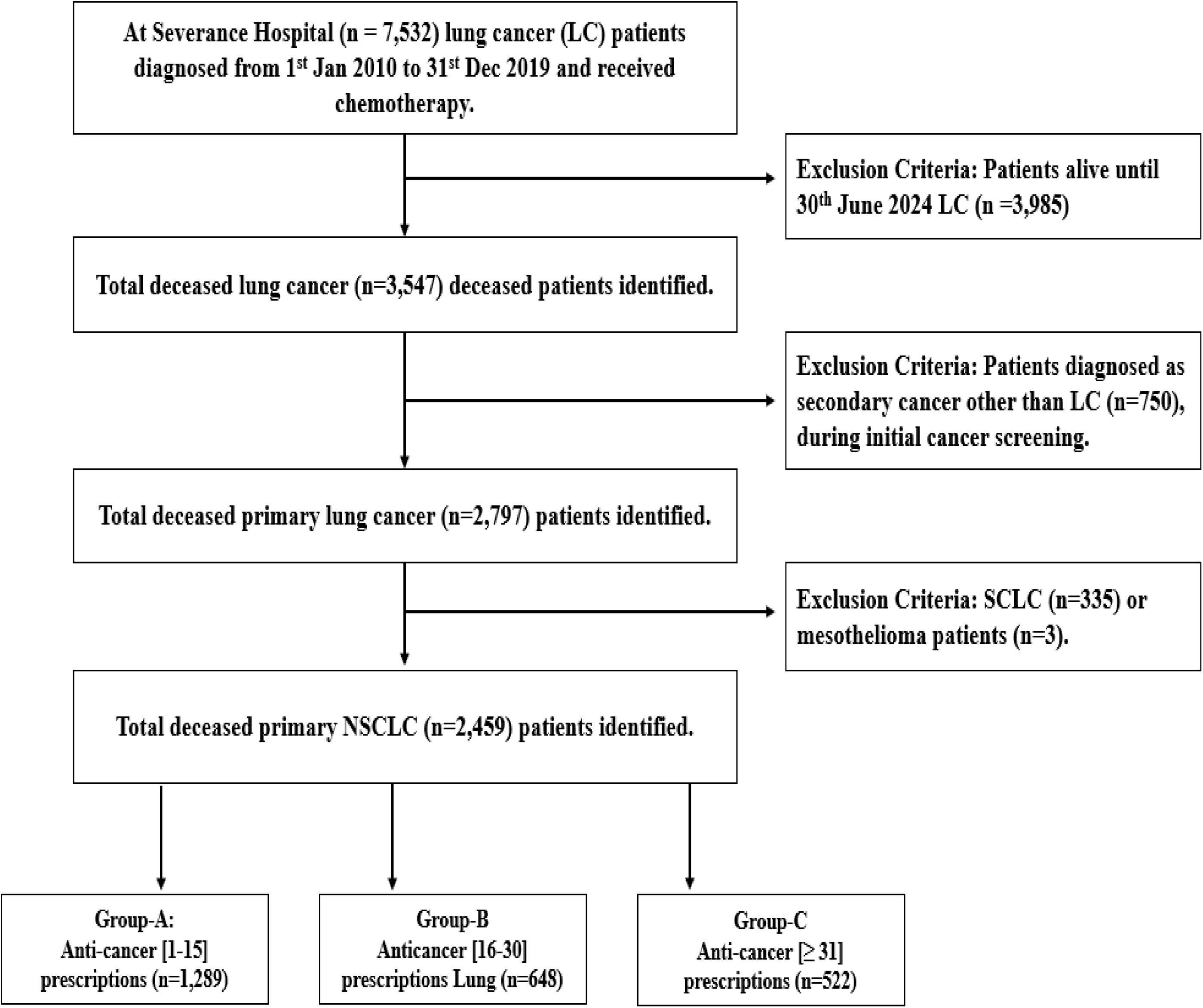
Flowchart for non-small cell lung cancer patient selection, patient population stratified based on anti-cancer drug prescription frequency.

### Stratification

This real-world study set up, intends to find anti-cancer drug response in NSCLC patients. Here patients were randomly stratified based on chemotherapy prescription frequency. Here groups “A”, “B”, and “C” represents patient population received treatment with various prescriptions of anti-cancer drugs [1–15], [16–30], and [31 or above] frequency respectively. Patient population was not stratified based on anti-cancer duration or anti-cancer total dosage as these two factors did not allocate patients as per patient’s inherent ability to withstand chemotherapy dose. Since patient’s dose selection was based on various factors such as age, gender, BSA, tumor location, size, and stage. Moreover, in this study 90% patients received chemotherapy as systemic formulation or injection. To investigate longitudinal chemotherapy response, prescription frequency is most suitable variable for stratification due to weekly or biweekly schedule of chemotherapy infusion and oral formulation are prescribed on daily or monthly basis. Previously, Xu et al. applied accurate but meticulous method to segregate patient population with chemotherapy tolerance or intolerance based on patients requiring chemo dose reduction, drug discontinuation or dose delay due to toxicities [33].

### Data collection

Lung cancer patients were searched in EMR using international classification of diseases-10 (ICD-10) code C34, information of gender, date of birth (DOB), surgery, treatment type, cancer stage, surgery, radiotherapy, comorbidity, performance score Eastern cooperative oncology group PS ECOG, NSCLC type, stage, BSA, weight, dates of surgery, radiotherapy, lab values of RBC, WBC, platelets, and albumin at baseline were retrieved. Complete information of radiological data, anti-cancer drug regimen, diagnosis dates of comorbidities, brand/generic names of anti-cancer drugs, and surgery were retrieved from EMR database. Comorbidities were searched with respective ICD-10 codes for acute/chronic kidney, heart, liver, thyroid, pulmonary diseases, and psychological disorders. Main variables considered as contributing factors of treatment response are age, gender, PS ECOG, BSA, surgery, radiotherapy, and comorbidity are shown as baseline characteristics in **Table 1**.

**Table 1:** Post stratification non-small cell lung cancer patient baseline characteristics. ^1^n (%); ^2^Mean ± SD (standard deviation).

| Characteristic | Overall<br>N = 2,459 | Group-A<br>N = 1,289 | Group-B<br>N = 648 | Group-C<br>N = 522 |
| --- | --- | --- | --- | --- |
| Gender<sup>1</sup> |  |  |  |  |
| Female | 779 (31.7) | 386 (29.9) | 202 (31.2) | 191 (36.6) |
| Male | 1,680 (68.3) | 903 (70.1) | 446 (68.8) | 331 (63.4) |
| Age<sup>1</sup> |  |  |  |  |
| Adult (≤64 years) | 1,288 (52.4) | 600 (46.5) | 365 (56.3) | 323 (61.9) |
| Aged (≥65 years) | 1,171 (47.6) | 689 (53.5) | 283 (43.7) | 199 (38.1) |
| ECOG<sup>1</sup> |  |  |  |  |
| ECOG 0 | 2,346 (95.4) | 1,218 (94.5) | 626 (96.6) | 502 (96.2) |
| ECOG ≥1 | 113 (4.6) | 71 (5.5) | 22 (3.4) | 20 (3.8) |
| NSCLC type<sup>1</sup> |  |  |  |  |
| Adenocarcinoma | 1,683 (68.4) | 878 (68.1) | 436 (67.3) | 369 (70.7) |
| Large-cell | 10 (0.4) | 5 (0.4) | 3 (0.5) | 2 (0.4) |
| Squamous cell | 398 (16.2) | 212 (16.4) | 108 (16.7) | 78 (14.9) |
| Unspecified | 368 (15.0) | 194 (15.1) | 101 (15.6) | 73 (14.0) |
| NSCLC Stage<sup>1</sup> |  |  |  |  |
| Stage I/II | 108 (4.4) | 42 (3.3) | 39 (6.0) | 27 (5.2) |
| Stage III/IV | 2,351 (95.6) | 1,247 (96.7) | 609 (94.0) | 495 (94.8) |
| Surgery<sup>1</sup> |  |  |  |  |
| No | 1,892 (76.9) | 1,048 (81.3) | 483 (74.5) | 361 (69.2) |
| Yes | 567 (23.1) | 241 (18.7) | 165 (25.5) | 161 (30.8) |
| Lobectomy | 266 (10.8) | 117 (9.1) | 80 (12.3) | 69 (13.2) |
| Lung transplant | 1 (0.0) | 1 (0.1) | 0 (0.0) | 0 (0.0) |
| Lymph node dissection | 317 (12.9) | 138 (10.7) | 95 (14.7) | 84 (16.1) |
| Pleurectomy | 1 (0.0) | 1 (0.1) | 0 (0.0) | 0 (0.0) |
| Pneumonectomy | 24 (1.0) | 10 (0.8) | 6 (0.9) | 8 (1.5) |
| Segmentectomy | 8 (0.3) | 5 (0.4) | 2 (0.3) | 1 (0.2) |
| Thoracotomy | 7 (0.3) | 3 (0.2) | 3 (0.5) | 1 (0.2) |
| Thymectomy | 1 (0.0) | 1 (0.1) | 0 (0.0) | 0 (0.0) |
| Wedge resection | 139 (5.7) | 66 (5.1) | 35 (5.4) | 38 (7.3) |
| Other procedures | 360 (14.6) | 157 (12.2) | 96 (14.8) | 107 (20.5) |
| Radiotherapy<sup>1</sup> |  |  |  |  |
| No | 1,552 (63.1) | 972 (75.4) | 353 (54.5) | 227 (43.5) |
| Yes | 907 (36.9) | 317 (24.6) | 295 (45.5) | 295 (56.5) |
| Comorbidity<sup>1</sup> |  |  |  |  |
| No | 1323 (53.8) | 747 (58.0) | 338 (52.2) | 238 (45.6) |
| Yes | 1136 (46.2) | 542 (42.0) | 310 (47.8) | 284 (54.4) |
| BSA<sup>2</sup> |  |  |  |  |
| ≤1.65 | 1,132 (46.0) | 642 (49.8) | 278 (42.9) | 212 (40.6) |
| ≥1.66 | 1,327 (54.0) | 647 (50.2) | 370 (57.1) | 310 (59.4) |
| Platelets (10x10<sup>12</sup>/L)<sup>2</sup> | 278.2 ± 112.4 | 282.1 ± 118.8 | 276.1 ± 108.5 | 271.0 ± 100.3 |
| RBC (10x10<sup>12</sup>/L)<sup>2</sup> | 4.4 ± 0.5 | 4.3 ± 0.6 | 4.4 ± 0.5 | 4.5 ± 0.5 |
| WBC (10x10<sup>3</sup> cells/ul)<sup>2</sup> | 8.4 ± 6.5 | 9.1 ± 6.5 | 7.9 ± 5.3 | 7.4 ± 7.8 |
| Albumin (g/dL)<sup>2</sup> | 3.6 ± 0.7 | 3.5 ± 0.7 | 3.7 ± 0.6 | 3.9 ± 0.6 |

### Outcome Measures

Overall survival was estimated as the interval between the date of first treatment to death and progression-free survival estimated using interval in diagnosis to date of progression or death. Post-diagnosis survival and post-chemotherapy end survival (in months), were also estimated based on mean interval period from date of diagnosis or end of treatment until death, respectively [34]. During chemotherapy metastasis was estimated from disease progression during overall period of chemotherapy. Metastatic site change was estimated from recurrence of new tumor after initial diagnosis within lung or other organs [21]. Patients of stratified groups were further divided as per treatment formulation type (injection, oral, & injection plus oral) and chemotherapy classes. Here, mean survival (months) post-treatment start to death were estimated in all analyses to identify drug response. Severe/non-severe hematological adverse events liver impairment and kidney failures as non-hematological events observed post-treatment (within 1-year) were reported as outcome.

### Prognosis of Treatment Response

To prognose anti-cancer drug response patients were further sorted as per resected or unresected status. Average WBC counts 7 days prior to drug initiation were obtained for analyses from EMR. A total of 152,790 WBC laboratory values for (n=2129/2459) patients are available, total 3,927 WBC lab values were 7-days pre-treatment WBC counts utilized for analysis. The main reason to use 7 days pre-treatment laboratory values because WBC count may vary beyond this time period in any cancer patient (35). This allowed to include (n=2129) patients with available pre-treatment WBC counts required for prognosis of treatment response.

### Statistical Analysis

Descriptive statistics were used to summarize patient characteristics and to evaluate treatment patterns in stratified sub-groups. The percentage of patients deceased at 1 month, 6 months, 1, 2, 3, 4, and 5 years after chemotherapy initiation was examined. Here 95% confidence interval (CI) around the coefficient estimate, as well as an associated *p*-value, were calculated using large sample approximations. Multivariate regression analysis was performed on variables such as age, gender, stage, BSA, PS ECOG, NSCLC type, surgery, radiotherapy, comorbidity, treatment type, and anti-cancer drugs. To identify odds ratio of 1-year mortality unadjusted statistical analyses of these variables were performed to find independent contributing factors for treatment response. Time-to-death (month) post treatment start was calculated for individual patients. Statistical difference in mean days survived was compared in both resected/unresected sub-groups among normal Vs leukopenia or leukocytosis in all stratified sub-groups. Response of anti-cancer agents mean overall survival (months) post-initiation of chemotherapy were applied in multivariate and Kaplan-Meier analysis for OS and PFS estimation. All analyses were performed in R-studio (ver. 3.6.1; R Foundation for Statistical Computing, Vienna, Austria).

## Results

### Baseline Characteristics

The baseline characteristics of 2459 NSCLC patients included in this study consisted of 779 female (31.7%) and 1680 male (68.3%). The median age of NSCLC patients were 64 years. At baseline performance status were ECOG 0, ECOG 1, and ECOG 2 in 95.4% (n=2346), 4.4% (n=108), 0.2% (n=5) patients, respectively. At baseline patients diagnosed with adenocarcinoma, large-cell, squamous cell carcinoma and unspecified in 68.4% (n=1683) patients, 0.4 (n=10), 16.2 (n=398), and 15% (n=368), respectively. Stage-I NSCLC was diagnosed in 16 (0.6%) patients, stage-II in 92 (3.7%) patients, stage-III in 474 (19.3%) patients, stage-IV in 1877 (76.2%) patients. In 567 (23.1%) patients were eligible for surgery, and 907 (36.9%) patients were recipient of radiotherapy, 1136 (46.2%) patients were found comorbid with various diseases such as diabetes, cardiovascular, liver and kidney diseases. Mean body weight was 61.8 kg, mean body surface area was 1.7 (m^2^). Mean RBC, WBC, platelets, neutrophil, and albumin were 4.4x10^12/L, 8.4x10^3 cells/ul, 278.2x10^12/L, 5.5x10^3 cells/ul and 3.6 g/dL, respectively. Pre/post stratification baseline characteristics with complete list of 55 variables are provided in supplementary file 1 (Table 1 - 2). Number of patients treated with singlet, doublet, triplet and quadruplet agents are presented in supplementary file 1 (Table 3 to 6). Various types of anti-cancer agents as per generic names, class and formulation type is available in supplementary file 1 (Table 8). Various types of surgeries considered as other procedures are listed in supplementary file 1 (Table 9).

**Table 2:**
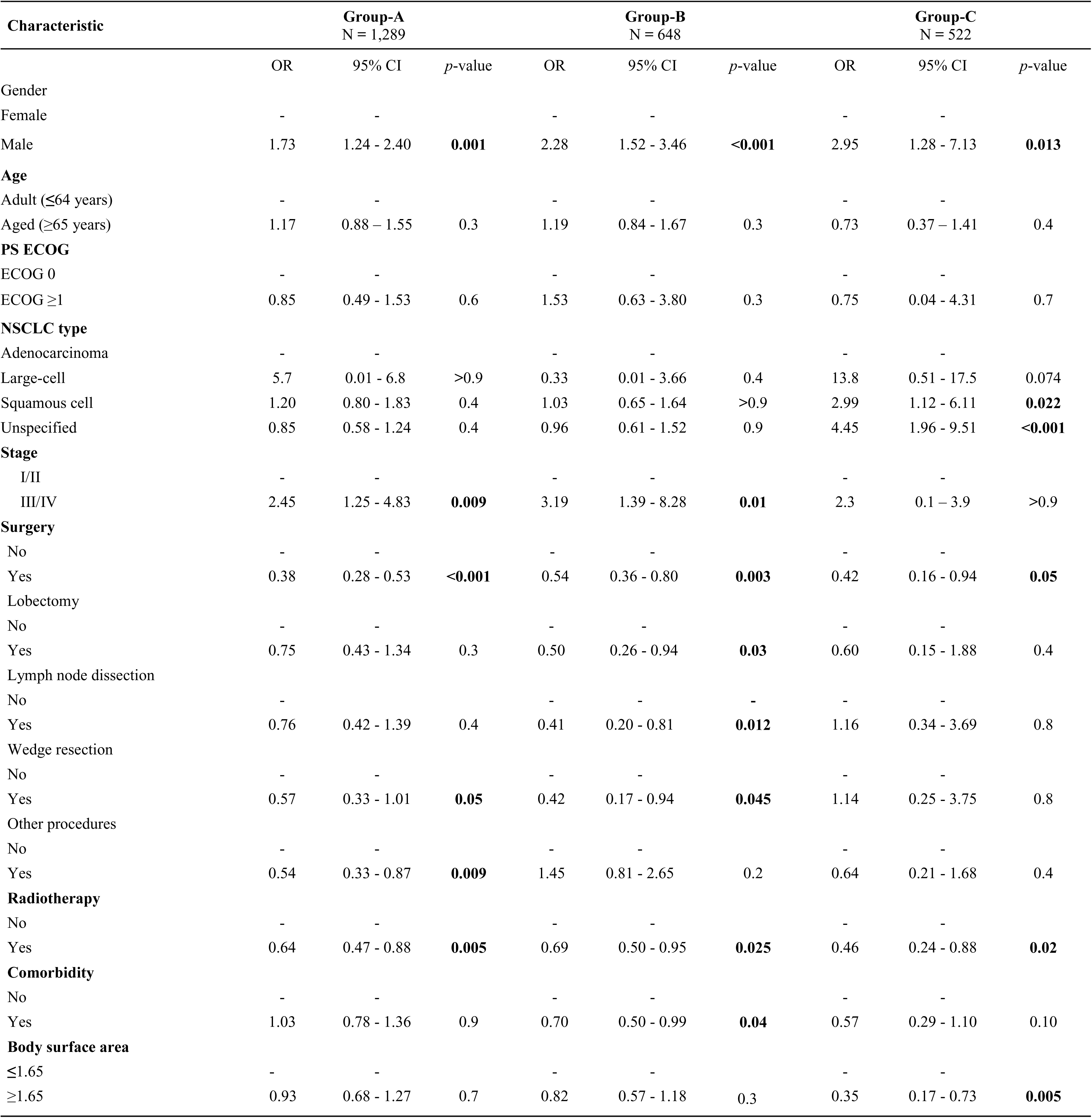
Odds of 1-year mortality in chemotherapy prescription frequency stratified sub-groups of non-small cell lung cancer. Abbreviations: CI; Confidence Interval, OR; Odds Ratio, NSCLC; non-small cell lung cancer, PS ECOG; Performance status Eastern cooperative Oncology Group.

**Table 3:**
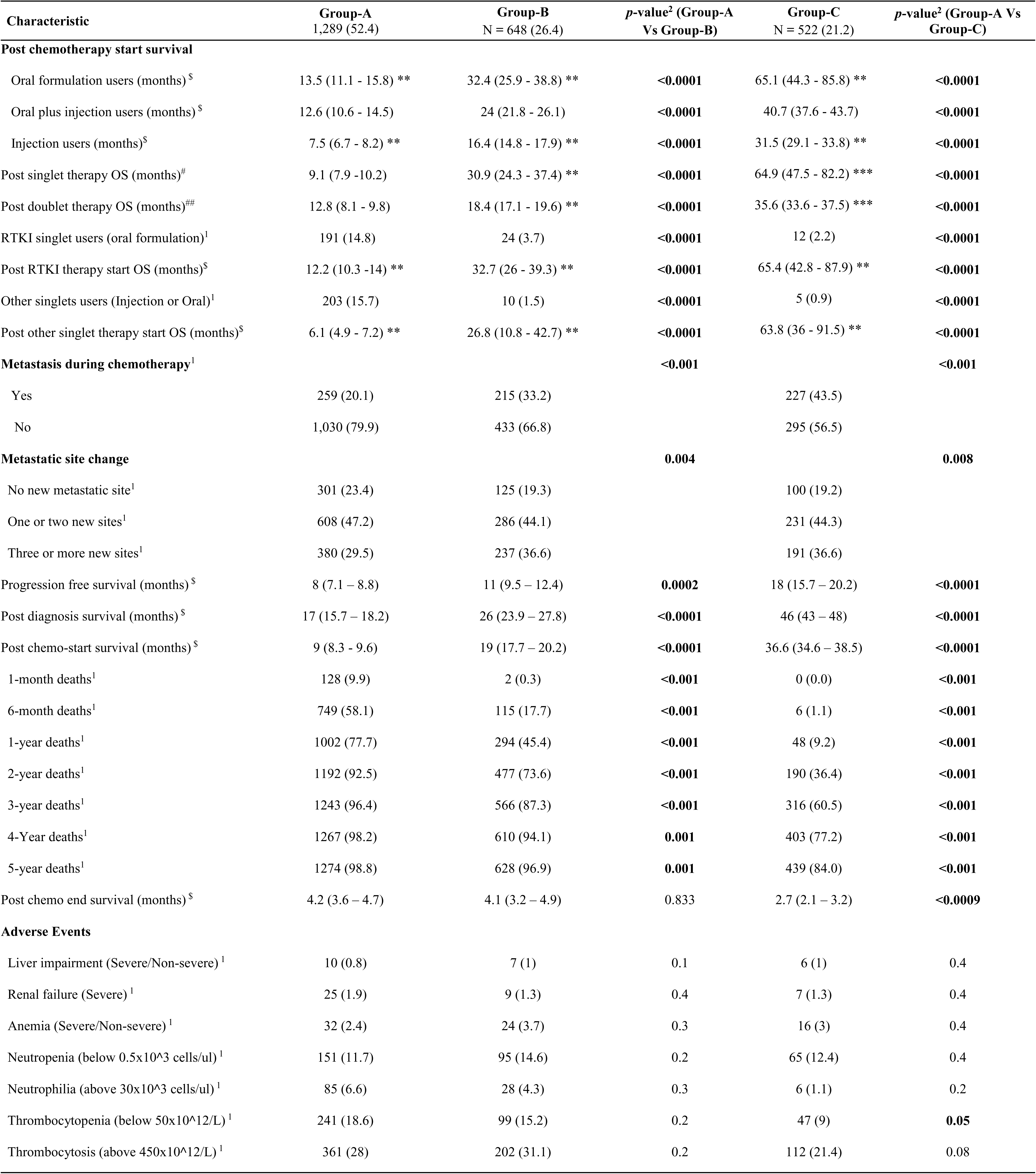
Outcome in chemotherapy prescription stratified sub-groups in non-small cell lung cancer (NSCLC) patients. ^1^n (%); number of patients, p-value; ^2^Pearson’s Chi-squared test. ^$^Mean, 95% confidence Interval (CI). Based on outcome group-A is considered as non-responder, group-B as moderate responder, and group-C as responder of anti-cancer drugs. ^#^(N=445) total singlet users, group-A (n=394), group-B (n=34), group-C (n=17). Total doublet users ^##^(N=2014), group-A (n=895), group-B (n=614), group-C (n=505). Among same group statistical significance (*p*-value) is shown using *** <0.0001; ** 0.001, and * 0.05. All adverse events occurred within 1-year post-chemotherapy initiation are shown above, patients experiencing hepatic/renal or hematological events prior to chemotherapy were excluded.

### Multivariate analysis of risk score

1-Year risk of mortality in males were higher compared to females across all stratified groups A, B, & C with significant statistical difference of *p*-value <0.0001; 0.001, and 0.05 respectively. There was no statistical difference in mortality risk among aged Vs adults, and ECOG 0 Vs ECOG ≥1 patients. In group-C patients diagnosed with squamous cell carcinoma and unspecified NSCLC were at statistically higher risk of 1-year mortality compared to adenocarcinoma patients with p-value (0.02 and <0.001). NSCLC stage I/II patients were at lower risk compared to stage III/IV patients in all three sub-groups with statistical significance of p-value (0.009, 0.01, >0.9). Radiotherapy recipients were at lower risk of mortality compared to radiotherapy untreated with statistical significance of *p*-value (0.005, 0.02, and 0.02). In group-B comorbid patients showed reduced risk of 1-year mortality with statistical significance of *p*-value 0.04. Patients in group-C patients with BSA ≥1.65 showed lesser risk of mortality compared to **≤**1.65 BSA patients with *p*-value 0.005. Resected patients were at lower risk of 1-year mortality compared to unresected with statistical significance in groups A, B, & C *p*-value <0.001, 0.003, 0.05, respectively. Wedge resection and other surgical procedures were beneficial in risk reduction compared to surgically non-intervened patients of group-A with *p*-value (0.05 and 0.009). Lobectomy, lymph node dissection, and wedge resection were beneficial for group-B patients compared to surgically unresected patients with *p*-value (0.03, 0.01, 0.04). Group-C resected Vs unresected patients showed no statistical difference in 1-year mortality risk. Results of multivariate analysis is available in **Table 2**.

### Treatment Outcomes

Metastasis during overall period of chemotherapy was found in 259/1289 (20.1%) patients of group-A, compared to 215/648 (33.2%) patients of group-B with statistical significance of *p*-value <0.001. In group-C 227/522 (43.5) patients were found to have metastasis with significant difference of *p*-value <0.001 compared to group-A. No new metastasis during therapy was highest in group-A (79.9%) compared to group-B (66.8%) or C (56.5%) patients. After initial diagnosis no new metastasis was found in (n=301) 23.4% patients of group-A, compared to group-B (n=125) 19.3% patients, and in group-C (n=100) 19.2% patients. One or two new metastatic sites in group-A (47.2%), compared to group-B (44.1%), and group-C (44.3%). In group-A 29.5% patients were diagnosed with three or more metastatic sites compared to group-B 36.6% patients, and group-C with 36.6% patients. Overall statistical significance of 0.004 was found in group-A Vs group-B and statistical significance of 0.008 in group-A Vs group-C. Based on these outcomes group-A is considered as non-responder, group-B as moderate responder, and group-C as responder of anti-cancer drugs. Post treatment initiation survival probability of NSCLC patients shown using Kaplan-Meier curve in **Figure 2**.

**Figure 2:**
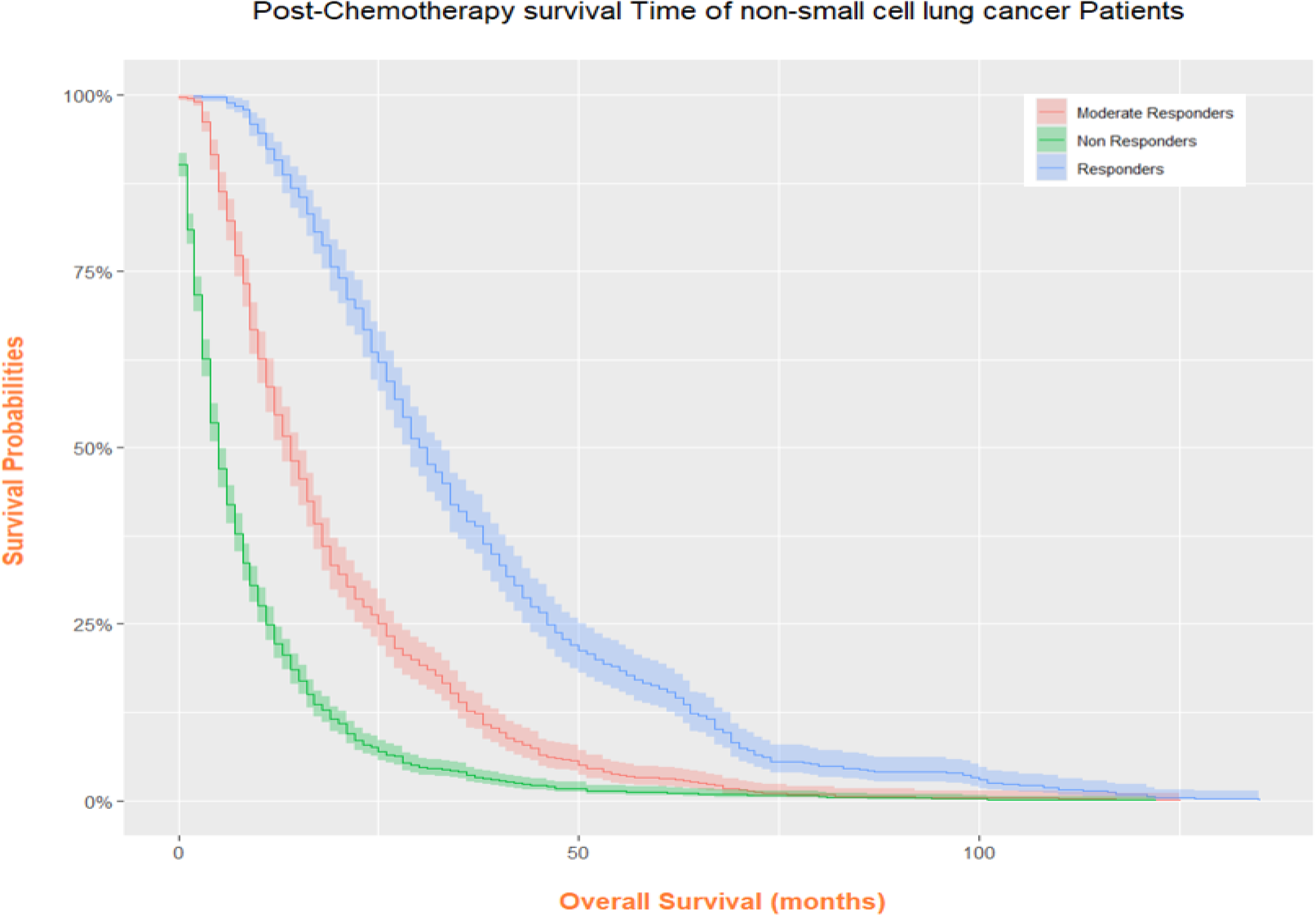
Overall survival post chemotherapy initiation in non-small cell lung cancer patients. Based on the outcome of treatment Group-A; Non-responder, Group-B; Moderate-responder, and Group-C; are Responder of treatment or chemotherapeutic regimen.

All survival analysis is reported as mean OS in months. Post-treatment initiation overall survival in group-A patients 9 months (95% 8.3 - 9.6), in group-B 19 months (95% CI 17.7 - 20.2) and in group-C 36.6 months (95% CI 34.6 - 38.5). Post-diagnosis overall survival in group-A patients were 17 months (95% CI 15.7 - 18.2), in group-B 26 months (95% CI 23.9 - 27.8), in group-C 46 months (95% CI 43 - 48). For both post treatment OS and post-diagnosis there is statistical significance of <0.0001 among group-A Vs B and group-A Vs C, respectively. PFS in group-A is 8 months (95% CI 7.1 - 8.8), group-B 11 months (95% CI 9.5 - 12.4) with statistical significance of *p*-value 0.0002, in group-C 18 months (95% CI 15.7 - 20.2) compared to group-A there is statistical significance of *p*-value <0.0001. Post treatment end OS is 4.2 months (3.6 – 4.7), in group-B 4.1 month (3.2 - 4.9) with no statistical significance among group-A Vs B p-value 0.833; and in group-C 2.7 months (2.1 - 3.2) with statistical difference of <0.0009. Mortality rate within 1-month in group-A was 9.9%, in group-B was 0.3% with statistical significance of *p*-value <0.001. There were no deaths in group-C. Mortality rate after 5 years of treatment was 98.8% in group-A, 96.9% in group-B *p*-value 0.001; and in group-C 84% with p-value <0.001. Survival analysis based on anti-cancer formulation type oral, injection, and injection plus oral were found to be significantly shorter in group-A compared to group-B or C patients. In group-A oral formulation users survived 13.5 months (11.1 - 15.8) compared to injection or systemic formulation users 7.5 months (6.7 - 8.2) with *p*-value 0.001. In total 10.3% patients survived more than five-years. Group-A (n=877) 68% patients died within 6 months whereas group-B (n=177) patients 18% died within 6 months, and in group-C (n=6) 1% patients died within 6 months. Total 1527/2459 (62%) patients were treated with first-line doublet combinations of alkylating/anti-mitotic, alkylating/Taxane, and antimetabolite/alkylating or with singlet agent oral RTKI drugs. Group-A patients were non-responsive toward first-line chemotherapeutic regimen or targeted therapy. After receiving doublets within 1-year mortality rate was highest in 79.2% group-A, compared to group-B 47% or group-C 9.3% patients. Group-A (21%) patients were prescribed with triplets and quadruplet chemo classes. Results of treatment outcome is provided in Table 3. Year wise mortality rate of (n=2014) doublet users is provided in Table 4. Year wise mortality rate as per treatment regimen is provided in supplementary file 1 (Table 7). Year wise mortality rate of patients treated with singlet, doublet, triplet and quadruplet agents are presented in supplementary file 2 (Figure 1 to 15). Pre/post lung cancer diagnosis associated year wise mortality rate in three patient groups is available in supplementary file 2 (Table 16).

**Table 4:** Year wise mortality rates in first-line doublet users ^1^(N=2014) diagnosed with NSCLC in stratified groups. ^1^n (%); number of patients. Refer supplementary Table 1-15 for detailed insight of mortalities in each group.

| Year wise Deaths<br>in groups | Overall<br>N=2014 | Alkylating,<br>Anti-mitotic | Alkylating,<br>Pyrimidines | Alkylating,<br>Taxane | Antimetabolite,<br>Alkylating | Antimetabolite,<br>Taxane | Other doublet<br>combinations | Topoisomerase<br>II Inhibitor,<br>Alkylating | RTKI,<br>Alkylating | RTKI,<br>Antimetabolite |
| --- | --- | --- | --- | --- | --- | --- | --- | --- | --- | --- |
| Group-A | 895 (44.4) | N = 46<sup>1</sup> | N = 57<sup>1</sup> | N = 359<sup>1</sup> | N = 221<sup>1</sup> | N = 19<sup>1</sup> | N = 102<sup>1</sup> | N = 36<sup>1</sup> | N = 14<sup>1</sup> | N = 41<sup>1</sup> |
| 1-Year Deaths | 709 (79.2) | 25 (54.3) | 52 (91.2) | 280 (78.0) | 194 (87.8) | 13 (68.4) | 82 (80.4) | 33 (91.7) | 11 (78.6) | 19 (46.3) |
| 2-Year Deaths | 121 (13.5) | 7 (15.2) | 2 (3.5) | 57 (15.9) | 19 (8.6) | 4 (21.1) | 14 (13.7) | 2 (5.6) | 0 (0.0) | 16 (39.0) |
| 3-Year Deaths | 35 (3.9) | 8 (17.4) | 1 (1.8) | 10 (2.8) | 5 (2.3) | 1 (5.3) | 4 (3.9) | 0 (0.0) | 0 (0.0) | 6 (14.6) |
| ≥4-Year Deaths | 30 (3.4) | 6 (13.0) | 2 (3.5) | 12 (3.3) | 3 (1.4) | 1 (5.3) | 2 (2.0) | 1 (2.8) | 3 (21.4) | 0 (0.0) |
| Group-B | 614 (30.4) | N = 64<sup>1</sup> | N = 57<sup>1</sup> | N = 191<sup>1</sup> | N = 150<sup>1</sup> | N = 4<sup>1</sup> | N = 45<sup>1</sup> | N = 52<sup>1</sup> | N = 21<sup>1</sup> | N = 30<sup>1</sup> |
| 1-Year Deaths | 289 (47.1) | 15 (23.4) | 44 (77.2) | 77 (40.3) | 87 (58.0) | 0 (0.0) | 18 (40.0) | 39 (75.0) | 1 (4.8) | 8 (26.7) |
| 2-Year Deaths | 174 (28.3) | 19 (29.7) | 11 (19.3) | 61 (31.9) | 34 (22.7) | 0 (0.0) | 17 (37.8) | 10 (19.2) | 9 (42.9) | 9 (30.0) |
| 3-Year Deaths | 82 (13.4) | 12 (18.8) | 1 (1.8) | 30 (15.7) | 17 (11.3) | 4 (100.0) | 7 (15.6) | 1 (1.9) | 4 (19.0) | 10 (33.3) |
| ≥4-Year Deaths | 69 (11.2) | 18 (28.1) | 1 (1.8) | 23 (12.0) | 12 (8.0) | 0 (0.0) | 3 (6.7) | 2 (3.8) | 7 (33.3) | 3 (10.0) |
| Group-C | 505 (25.0) | N = 50<sup>1</sup> | N = 39<sup>1</sup> | N = 120<sup>1</sup> | N = 99<sup>1</sup> | N = 11<sup>1</sup> | N = 68<sup>1</sup> | N = 33<sup>1</sup> | N = 43<sup>1</sup> | N = 42<sup>1</sup> |
| 1-Year Deaths | 47 (9.3) | 0 (0.0) | 8 (20.5) | 12 (10.0) | 11 (11.1) | 2 (18.2) | 4 (5.9) | 6 (18.2) | 3 (7.0) | 1 (2.4) |
| 2-Year Deaths | 140 (27.7) | 7 (14.0) | 17 (43.6) | 27 (22.5) | 36 (36.4) | 4 (36.4) | 20 (29.4) | 13 (39.4) | 9 (20.9) | 7 (16.7) |
| 3-Year Deaths | 124 (24.6) | 13 (26.0) | 6 (15.4) | 33 (27.5) | 20 (20.2) | 3 (27.3) | 22 (32.4) | 8 (24.2) | 7 (16.3) | 12 (28.6) |
| ≥4-Year Deaths | 194 (38.4) | 30 (60.0) | 8 (20.5) | 48 (40.0) | 32 (32.3) | 2 (18.2) | 22 (32.4) | 6 (18.2) | 24 (55.8) | 22 (52.4) |

### Prognosis of Chemotherapy response

Among surgically resected group-A, group-B, and group-C there is no significant survival difference in patients diagnosed with leukopenia and three sub-groups of normal WBC counts. Pre-chemotherapy WBC count (4 – 5.9 x 10^3^) diagnosed in 47 patients survived 19.5 months (95% CI 13 - 25.9) compared to 23 patients diagnosed with leukocytosis (≥10.8 x 10^3^) survived 5.3 months (95% CI 2.6 – 7.9) with *p*-value 0.004. Similarly, in unresected group-A and group-B patients, there is no major difference in survival post treatment. Pre-chemotherapy WBC count (4 – 5.9 x 10^3^) diagnosed in 138 patients survived 9.6 months (95% CI 7.9 – 11.2) compared to 243 patients diagnosed with leukocytosis (≥10.8 x 10^3^) survived 4.7 months (95% CI 3.9 – 5.4) with *p*-value <0.0001. In unresected group-C pre-chemotherapy WBC count (6 – 7.99 x 10^3^) and (8.1 – 10.80 x 10^3^) sub-groups survived significantly shorter compared to leukopenia sub-group patients with *p*-value 0.002 and 0.01 respectively. Table 4 is provided with sample size, OS mean, ±SD and (95% CI). Leukocytosis was found in (43%) patients died within 1 month significantly more compared to normal or leukopenia in NSCLC patients. In 21/227 (9.2%) RTKI oral formulation users survived 4.2 months (95% CI 0.9 – 7.4) significantly less compared to normal patients (n=67) survived 16 months (95% CI 11.6 – 20.5) *p*-value (0.005). Results of prognosis of chemotherapy response in non-small cell lung cancer (n=2129/2459) patients is available in Table 5.

**Table 5:** Prognosis of chemotherapy response based on average WBC counts obtained 7 days prior to chemotherapy initiation in non-small cell lung cancer (n=2129/2459) patients. Comparison of overall survival between leukopenia (>3.9 x 10^9^ cells/L or less) Vs normal (4-5.9x10^9^ cells/L) range of WBC counts, and leukocytosis (10.81 x 10^9^ cells/L or above) Vs normal (4-5.9x10^9^ cells/L) range of WBC counts.

|  | WBC Count |  | Group-A<br>N=1,136 |  |  | Group-B<br>N=582 |  |  |  | Group-C<br>N=411 |  |  |  |
| --- | --- | --- | --- | --- | --- | --- | --- | --- | --- | --- | --- | --- | --- |
| Resected (n=493) | WBC Range | N | OS (95%CI) | ±SD | p-value | N | OS (95%CI) | ±SD | p-value | N | OS (95% CI) | ±SD | p-value |
|  | ≤ 3.9 x 10<sup>9</sup> (a) | 9 | 14.7 (10.9 -18.4) | ±5.8 | (a Vs b) 0.5 | 7 | 24.2 (12.3 - 36) | ±16 | (a Vs b) 0.5 | 7 | 49 (25.3 – 72.7) | ±32.09 | (a Vs b) 0.9 |
|  | 4 - 5.9 x 10<sup>9</sup> (b) | 47 | 19.5 (13 - 25.9) | ±22.7 | (b Vs a) 0.5 | 37 | 28.2 (23.4 - 32.9) | ±14.8 | (b Vs a) 0.5 | 28 | 50.3 (38.8 – 61.7) | ±31.02 | (b Vs a) 0.9 |
|  | 6 - 7.9 x 10<sup>9</sup> (c) | 72 | 16.1 (11.2 - 20.9) | ±21.03 | (c Vs a) 0.8 | 55 | 27.8 (21.3 - 34.2) | ±24.5 | (c Vs a) 0.7 | 58 | 37.3 (32.6 – 41.9) | ±17.9 | (c Vs a) 0.1 |
|  | 8.1 - 10.80 x 10<sup>9</sup> (d) | 56 | 14.7 (9.1 - 20.2) | ±21.3 | (d Vs a) 0.1 | 39 | 24.4 (17.3 - 31.4) | ±22.6 | (d Vs a) 0.9 | 32 | 45.5 (35.3 – 55.6) | ±29.4 | (d Vs a) 0.7 |
|  | ≥10.81 x 10<sup>9</sup> (e) | 23 | 5.3 (2.6 - 7.9) | ±6.5 | (e Vs b) **0.004** | 11 | 18.9 (11.6 - 26.1) | ±12.2 | (e Vs b) 0.06 | 12 | 40.8 (26.3 – 55.2) | ±25.5 | (e Vs b) 0.3 |
| Unresected (n=1636) | WBC Range | N | OS (95%CI) | ±SD | p-value | N | OS (95%CI) | ±SD | p-value | N | OS (95% CI) | ±SD | p-value |
|  | ≤ 3.9 x 10<sup>9</sup> (f) | 36 | 7.8 (4.7 - 10.8) | ±9.3 | (f Vs g) 0.3 | 17 | 14.2 (9.2 - 19.1) | ±10.5 | (f Vs g) 0.2 | 10 | 52.1 (37.1 - 67) | ±24.2 | (f Vs g) 0.1 |
|  | 4 - 5.9 x 10<sup>9</sup> (g) | 138 | 9.6 (7.9 - 11.2) | ±9.9 | (g Vs f) 0.3 | 76 | 18.6 (15.4 - 21.7) | ±13.9 | (g Vs f) 0.2 | 56 | 37.1 (30.1 - 44) | ±26.5 | (g Vs f) 0.1 |
|  | 6 - 7.9 x 10<sup>9</sup> (h) | 240 | 8.5 (7.4 - 9.5) | ±8.6 | (h Vs f) 0.6 | 143 | 19 (16.4 -21.5) | ±15.7 | (h Vs f) 0.2 | 107 | 31.6 (27.8 - 35.3) | ±19.7 | (h Vs f) **0.002** |
|  | 8.1 - 10.80 x 10<sup>9</sup> (i) | 272 | 7.1 (5.7 - 8.4) | ±11.1 | (i Vs f) 0.7 | 114 | 15.8 (13.6 - 17.9) | ±11.9 | (i Vs f) 0.6 | 69 | 33.2 (28.1 - 38.2) | ±21.4 | (i Vs f) **0.01** |
|  | ≥10.81 x 10<sup>9</sup> (j) | 243 | 4.7 (3.9 - 5.4) | ±5.8 | (j Vs g) **<0.0001** | 83 | 12.2 (9.4 - 14.9) | ±13 | (j Vs g) **0.04** | 32 | 30.1 (24.1 - 36) | ±17.1 | (j Vs g) 0.1 |

## Discussion

Stratifying patient population based on tumor response previously failed to obtain statistical correlation between tumor response and other variables, survival was corelated to tumor response demonstrated using multivariate analysis in gastric cancer patients [21]. In this study it is shown for the first time significance of stratifying patient population as per chemotherapy prescriptions to identify chemotherapy response. Although screening of tumor from PET/CT scan reports after few cycles of chemotherapy is important and has remained a common clinical practice for identifying drug response.

Group-A patients were ineligible for intensive doses of chemotherapy or surgery after close examination during screening by healthcare professional. Group-A patients were withdrawn from treatment; drugs were discontinued due to toxicities arising from low doses and frequency of anti-cancer drugs causing disease progression and deaths [33]. Group-A patients prescribed with triplets and quadruplet chemo classes, frequently drugs were withdrawn or changed due to chemotherapy intolerance and toxicities. Maximum patients died within 1 month, 6 month and 1-year post treatment initiation in group-A compared to other two groups. Selection of safest drugs with high treatment dosage may result in fatality within a short duration [36]. Group-A patients were non-responsive toward first-line chemotherapeutic regimen, immunotherapies or targeted therapy compared to other groups. Comorbidities in group-A and C were associated with risk of 1-year mortality, previously comorbidity reportedly reduced quality-of-life in cancer patients [37–38]. Importance of surgical intervention was lowest in group-C compared to Group-A or B patients as patients responded well toward high doses of chemotherapy despite of numerous new metastases during ongoing chemotherapy regimen. Low BSA (**≤** 1.65 m^2^) prohibited group-A patients from receiving maximum dosage of chemotherapy for control of tumor growth [39]. Group-A patients were more dependent on other surgical procedures than Group-B or C patients are directly corelated to drug response. Based on these findings group-A patients are considered as non-responder of chemotherapy. Group-A patients were intolerant and resistant to chemotherapeutic regimen showed non-responsive toward anti-cancer agents.

Cancer patients non-responsive towards chemotherapy could be due to several reasons. Chemo-resistance could be due to mutation in drug targets, altered drug metabolism due to gene polymorphism, enhanced DNA repair mechanism of tumor cells, overexpression of P-glycoprotein (P-gp) in tumor cell promotes excessive anti-cancer drug elimination [40]. Chemo-resistance is the leading cause of deaths in NSCLC patients of this study. Group-A patients developed early chemo-resistance post-treatment initiation compared to other two groups, standard recommended treatment strategies were not beneficial for these NSCLC patients [41]. Chemotherapy intolerance was corelated to anti-cancer drug induced toxicities, hematological, other severe and life-threatening adverse events as well as renal insufficiency are potential reasons of mortality and nevertheless cancer progression due to drug withdrawal in majority of patients of all groups [33–42].

Recently, Roviello et al. in a retrospective study reported that prognosing patients non-responsive toward chemotherapy is of great importance. WBC counts are mainly used clinically to identify underlying risk of infections. In this study 19% patients were diagnosed with pre-treatment leukocytosis. Mainly leukocytosis patients should be closely monitored selected for safe dosing and treatment regimen mainly in unresected patients. Based on assimilated results it is recommended that WBC count prior to chemotherapy start could identify patients with highest risk of early mortality. Obtaining lab reports of WBC counts are cheaper than obtaining lymphocyte counts. Moreover, obtaining lab tests of blood cellular sub-types requires substantially more amount of blood withdrawal for analysis, it could be a challenge for patients comorbid with anemia.

Previously in ovarian cancer patients WBCs obtained 7 days pre-treatment showed leukopenia with survival benefit compared to normal WBC counts, there were no patients diagnosed with pre-treatment leukocytosis [35]. In another study pre-treatment WBC count above >7 was found to increased risk of survival, compared to below <7 WBC count in lung cancer patients and pre-treatment leukocytosis as risk was not shown [31]. Elevated total WBC counts in cancer patients was found to be a risk factor of early mortality in overall cancer patients [43]. Therefore, in this study it is shown for the first time prognostic significance of leukocytosis as major risk factor for survival in NSCLC patients.

At present chemotherapy cessation in responder is another challenge because these patients died shortly after end or during ongoing chemotherapy. It is relatively important to identify patient’s ineligibility for higher doses of chemotherapy as well as chemotherapy cessation in patients undergoing prolonged treatment, as till-date no biomarkers are available to suggest therapy cessation [44]. Immunosuppressants such as everolimus and methotrexate were not as beneficial as pemetrexed (anti-metabolite) or vinorelbine (anti-mitotic) for NSCLC treatment. Pemetrexed and vinorelbine due to its dual mode of action as immunosuppressant and tumor growth inhibitor were found to be better treatment drugs used along with cisplatin/carboplatin. Combination of pemetrexed plus cisplatin or vinorelbine plus cisplatin are commonly used standard comparator drugs in clinical trials of NSCLC [45]. In this study immunotherapy (biologics) prescribed such as trastuzumab, bevacizumab, durvalumab, ramucirumab, ipilimumab, rituximab, and cetuximab were not beneficial compared to RTKI in NSCLC patients. Immunotherapy virotherapy cotreatment regimen against various cancers earlier demonstrated advanced tumor clearance [46].

The main limitations of this study are patient’s economic background, smoking, drinking, and health insurance status are important confounding factors were not taken into consideration in this study. Current smoking status could be an independent contributing factor for mortality in lung cancer patients not considered in this study [47]. In this study more leverages were applied on drug response stratification or chemo type/formulation, comorbidities, surgeries, and radiotherapy rather than smoking as confounding factors, also suggested by Myers et al. [48]. Earlier in a cohort study without patient group stratification found smoking to be directly related with low survival rate in NSCLC patients [24]. Due to low sample size of kinase inhibitors (oral) users, RTKI remains the best drug among all classes could be a potential bias. Two patients in group-B died within 1 month received more than 15 prescriptions of daily oral RTKI, usually RTKI are prescribed to be ingested at home and for majority of patients oral formulations are prescribed on monthly basis. No additional measures during analysis were taken to separate these two patients could be considered as a limitation and inaccuracy of study design.

A future prospective study could be conducted by pretreating patients with least toxic anti-inflammatory drugs such as dexamethasone 4mg twice daily for short duration at least three to seven days prior to therapy initiation in NSCLC patients diagnosed with pre-treatment leukocytosis [45]. Using anti-inflammatory drugs prior to cancer treatment could be beneficial to substantially reduce early activation of specific subset of t-lymphocytes responsible for aggressive and selective killing of tumor cells. Reducing chemotactic movement of armed t-lymphocytes against solid tumors may significantly enhance clinical efficacy of anti-cancer agents [49]. Often at the end or beginning of chemotherapy patients with neutropenia are treated with G-CSF to maximize efficacy of chemotherapy and to prolong survival [31].

## Conclusion

Similar investigational study design could be applied in any cancer patients would lead to same findings is a major claim of this study. Pre-treatment leukocytosis was related to poor survival in NSCLC patients selected for mild or moderate dosage of chemotherapy regimen irrespective of surgery status. Patients selected for heavy doses of chemotherapy should also be closely monitered for leukocytosis. Patients with no surgical interventions were more vulnerable to early mortality compared to surgically intervened responder of chemotherapy. However, it was found that chemotherapy responders are less prone to 1-year mortality irrespective of surgery status or comorbid condition. Oral formulation of RTKI was beneficial in all groups of NSCLC patients, RTKI was ineffective in patients with pre-treatment leukocytosis. Cumulative 5-year survival rate was 10.3% in NSCLC patients consistent with previous reports. RTKI usage was safe and beneficial compared to chemotherapeutic agents, due to selective mutation status and high cost of RTKI often not prescribed to patients of low or medium-income backgrounds. Based on assimilated results it is clear that chemotherapeutic doses recommended by manufacturers are unsafe for chemotherapy non-responders, minimum safe dose (MSD) must be revised for chemotherapy non-responders. Low-dose metronomic (LDM) chemotherapy could be beneficial for intolerant or drug-resistance toward standard treatment dose.

## Supporting information

STROBE_Checklist

## Funding

No funding was received to assist with the preparation of this manuscript.

## Competing Interest Statement

The author has no competing interest to declare.

## Data availability

Data will be made available upon request, post-approval from Yonsei University IRB.

## Author Declaration

I hereby confirm that all relevant ethical guidelines have been followed, and any necessary IRB and/or ethics committee approval was obtained. The studies involving human participants were waived by the Institutional review board (IRB) of Yonsei University because patient identity was masked during data retrieval. Written informed consent form for participation was not required for this study in accordance with the Korean national legislation and the institutional requirements.

## Acknowledgement

The author would like to thank professor Kyungsoo park (Department of pharmacology, Yonsei University) for providing access to severance hospital database for obtaining patient’s electronic health records.

## Notes

### Competing Interest Statement

The authors have declared no competing interest.

