## Supplementary File 1 for "Prognostic Features of Anti-Cancer Drugs Response in Resected/Unresected Primary Non-Small Cell Lung Cancer: A Retrospective Cohort Study"

Saikat Samadder^1*^

^1^Department of Pharmacology, Yonsei University College of Medicine, 50 Yonsei-ro, Seodaemun-Gu, Seoul, Korea

^*^ Correspondence: Saikat Samadder

**Supplementary file 1**

**Table of contents**

1. Supplementary table 1: Pre-stratification NSCLC (N=2459) patient characteristics. ---------------------------------------------- Page [1-6]

2. Supplementary table 2: Post-stratification NSCLC (N=2459) patient characteristics. ------------------------------------------- Page [7-10]

3. Supplementary table 3: List of singlet chemo classes applied on NSCLC (n=445) patients -------------------------------------- Page [11]

4. Supplementary table 4: List of various doublet agents applied on NSCLC (n=822) patients. ------------------------------------ Page [12]

5. Supplementary table 5: List of various triplet agents applied on NSCLC (n=623) patients. -------------------------------------- Page [13]

6. Supplementary table 6: List of various quadruplet agents applied on NSCLC (n=569) patients. -------------------------------- Page [14]

7. Supplementary table 7: Year wise deaths as per cancer treatment type received by NSCLC (n=2459) patients --------------- Page [15]

8. Supplementary table 8: List of generic names, chemo-class, and formulation type of drugs prescribed to NSCLC patient -- Page [16]

9. Supplementary table 9: List of 158 surgeries considered as other procedures ---------------------------------------------------- Page [17-19]

| **Characteristic** | **Lung cancer**  N = 2,459^1^ |
| --- | --- |
| Gender |  |
| Female | 779 (31.7) |
| Male | 1,680 (68.3) |
| Age (mean ±SD)  Age (median) | 63.1 ± 10.9  64 |
| Performance status ECOG |  |
| 0 | 2,346 (95.4) |
| 1 | 108 (4.4) |
| 2 | 5 (0.2) |
| NSCLC type |  |
| Adenocarcinoma | 1,683 (68.4) |
| Large-cell | 10 (0.4) |
| Squamous cell | 398 (16.2) |
| Unspecified | 368 (15.0) |
| Stage |  |
| I | 6 (0.2) |
| IA | 1 (0.0) |
| IB | 9 (0.4) |
| II | 25 (1.0) |
| IIA | 29 (1.2) |
| IIB | 38 (1.5) |
| III | 154 (6.3) |
| IIIA | 203 (8.3) |
| IIIB | 116 (4.7) |
| IIIC | 1 (0.0) |
| IV | 1,752 (71.2) |
| IVA | 6 (0.2) |
| IVB | 6 (0.2) |
| Recurred  Adjuvant  Consolidation  Induction  Maintenance  Neoadjuvant  Palliative  Salvage | 113 (4.6)  200 (7.7)  6 (0.2)  23 (0.9)  2 (0.1)  94 (3.8)  2057 (83.6)  77 (3.1) |
| Surgery |  |
| No | 1,892 (76.9) |
| Yes | 567 (23.1) |
| Radiotherapy |  |
| No | 1,552 (63.1) |
| Yes | 907 (36.9) |
| Weight (kg) | 61.8 ± 10.5 |
| Platelets (10x10^12/L) | 278.2 ± 112.4 |
| RBC (10x10^12/L) | 4.4 ± 0.5 |
| WBC (10x10^3 cells/ul) | 8.4 ± 6.5 |
| Neutrophil (10x10^3 cells/ul)  Albumin (g/dL) | 5.5 ± 2.9  3.6 ± 0.7 |
| Body surface area | 1.7 ± 0.2 |
| Chemo duration | 422.4 ± 544.9 |
| Chemo dosage | 22,008.2 ± 74,070.6 |
| Chemotherapy prescriptions | 21.4 ± 24.1 |
| Formulation type |  |
| Injection users | 1,642 (66.8) |
| Oral users | 234 (9.5) |
| Injection & Oral users | 583 (23.7) |
| Metastatic site change |  |
| 0 | 526 (21.4) |
| 1 | 627 (25.5) |
| 2 | 498 (20.3) |
| 3 | 396 (16.1) |
| 4 | 226 (9.2) |
| 5 | 105 (4.3) |
| 6 | 56 (2.3) |
| 7 | 19 (0.8) |
| 8 | 4 (0.2) |
| 9 | 2 (0.1) |
| During chemotherapy metastasis |  |
| No | 1,758 (71.5) |
| Yes | 701 (28.5) |
| Alkylating |  |
| No | 624 (25.4) |
| Yes | 1,835 (74.6) |
| Antiandrogen |  |
| No | 2,458 (100.0) |
| Yes | 1 (0.0) |
| Antimetabolite |  |
| No | 1,041 (42.3) |
| Yes | 1,418 (57.7) |
| Anti-mitotic |  |
| No | 2,080 (84.6) |
| Yes | 379 (15.4) |
| Biologics |  |
| No | 1,924 (78.2) |
| Yes | 535 (21.8) |
| Immunosuppressant |  |
| No | 2,439 (99.2) |
| Yes | 20 (0.8) |
| Kinase inhibitor |  |
| No | 2,449 (99.6) |
| Yes | 10 (0.4) |
| Pyrimidines |  |
| No | 1,855 (75.4) |
| Yes | 604 (24.6) |
| Receptor tyrosine kinase inhibitor |  |
| No | 1,667 (67.8) |
| Yes | 792 (32.2) |
| Taxane |  |
| No | 1,385 (56.3) |
| Yes | 1,074 (43.7) |
| Topoisomerase I/II inhibitor |  |
| No | 2,248 (91.4) |
| Yes | 211 (8.6) |
| **Chemo class** |  |
| Double classes | 822 (33.4) |
| Quadruple classes | 335 (13.6) |
| Quintuple or above classes | 234 (9.5) |
| Single class | 445 (18.1) |
| Comorbidity |  |
| No | 1,323 (53.8) |
| Yes | 1,136 (46.2) |
| Diabetes |  |
| No | 2,063 (83.9) |
| Yes | 396 (16.1) |
| Anemia |  |
| No | 2,293 (93.2) |
| Yes | 166 (6.8) |
| Fatty liver |  |
| No | 2,438 (99.1) |
| Yes | 21 (0.9) |
| Chronic liver disease |  |
| No | 2,402 (97.7) |
| Yes | 57 (2.3) |
| Anxiety |  |
| No | 2,266 (92.2) |
| Yes | 193 (7.8) |
| Obsessive compulsive disorder |  |
| No | 2,457 (99.9) |
| Yes | 2 (0.1) |
| Panic disorder |  |
| No | 2,456 (99.9) |
| Yes | 3 (0.1) |
| Chronic obstructive pulmonary disease |  |
| No | 2,076 (84.4) |
| Yes | 383 (15.6) |
| Chronic kidney disease |  |
| No | 2,392 (97.3) |
| Yes | 67 (2.7) |
| Renal failure |  |
| No | 2,344 (95.3) |
| Yes | 115 (4.7) |
| Coronary artery disease |  |
| No | 2,294 (93.3) |
| Yes | 165 (6.7) |
| Hypothyroidism |  |
| No | 2,331 (94.8) |
| Yes | 128 (5.2) |
| Hyperthyroidism |  |
| No | 2,429 (98.8) |
| Yes | 30 (1.2) |
| Lobectomy |  |
| No | 2,193 (89.2) |
| Yes | 266 (10.8) |
| Lung transplant |  |
| No | 2,458 (100.0) |
| Yes | 1 (0.0) |
| Lymph node dissection |  |
| No | 2,142 (87.1) |
| Yes | 317 (12.9) |
| Pleurectomy |  |
| No | 2,458 (100.0) |
| Yes | 1 (0.0) |
| Pneumonectomy |  |
| No | 2,435 (99.0) |
| Yes | 24 (1.0) |
| Segmentectomy |  |
| No | 2,451 (99.7) |
| Yes | 8 (0.3) |
| Thoracotomy |  |
| No | 2,452 (99.7) |
| Yes | 7 (0.3) |
| Thymectomy |  |
| No | 2,458 (100.0) |
| Yes | 1 (0.0) |
| Other procedure |  |
| No | 2,099 (85.4) |
| Yes | 360 (14.6) |
| Wedge resection |  |
| No | 2,320 (94.3) |
| Yes | 139 (5.7) |
| ^1^n (%); Mean ± SD | |

**Supplementary table 1:** Pre-stratification NSCLC (N=2459) patient characteristics.

| **Characteristic** | **Overall**  N = 2,459^1^ | **Group-A**  N = 1,289^1^ | **Group-B**  N = 648^1^ | **Group-C** N = 522^1^ |
| --- | --- | --- | --- | --- |
| **Gender** |  |  |  |  |
| Female | 779 (31.7) | 386 (29.9) | 202 (31.2) | 191 (36.6) |
| Male | 1,680 (68.3) | 903 (70.1) | 446 (68.8) | 331 (63.4) |
| **Age** | 63.1 ± 10.9 | 64.6 ± 11.0 | 62.4 ± 10.3 | 60.2 ± 11.0 |
| **PS ECOG** |  |  |  |  |
| 0 | 2,346 (95.4) | 1,218 (94.5) | 626 (96.6) | 502 (96.2) |
| 1 | 108 (4.4) | 66 (5.1) | 22 (3.4) | 20 (3.8) |
| 2 | 5 (0.2) | 5 (0.4) | 0 (0.0) | 0 (0.0) |
| **Non-small cell lung cancer type** |  |  |  |  |
| Adenocarcinoma | 1,683 (68.4) | 878 (68.1) | 436 (67.3) | 369 (70.7) |
| Large-cell | 10 (0.4) | 5 (0.4) | 3 (0.5) | 2 (0.4) |
| Squamous cell | 398 (16.2) | 212 (16.4) | 108 (16.7) | 78 (14.9) |
| Unspecified | 368 (15.0) | 194 (15.1) | 101 (15.6) | 73 (14.0) |
| **Stage** |  |  |  |  |
| I | 6 (0.2) | 1 (0.1) | 5 (0.8) | 0 (0.0) |
| IA | 1 (0.0) | 0 (0.0) | 1 (0.2) | 0 (0.0) |
| IB | 9 (0.4) | 5 (0.4) | 2 (0.3) | 2 (0.4) |
| II | 25 (1.0) | 13 (1.0) | 2 (0.3) | 10 (1.9) |
| IIA | 29 (1.2) | 9 (0.7) | 14 (2.2) | 6 (1.1) |
| IIB | 38 (1.5) | 14 (1.1) | 15 (2.3) | 9 (1.7) |
| III | 154 (6.3) | 67 (5.2) | 48 (7.4) | 39 (7.5) |
| IIIA | 203 (8.3) | 72 (5.6) | 71 (11.0) | 60 (11.5) |
| IIIB | 116 (4.7) | 42 (3.3) | 42 (6.5) | 32 (6.1) |
| IIIC | 1 (0.0) | 1 (0.1) | 0 (0.0) | 0 (0.0) |
| IV | 1,752 (71.2) | 1,008 (78.2) | 413 (63.7) | 331 (63.4) |
| IVA | 6 (0.2) | 4 (0.3) | 1 (0.2) | 1 (0.2) |
| IVB | 6 (0.2) | 3 (0.2) | 1 (0.2) | 2 (0.4) |
| Recurred | 113 (4.6) | 50 (3.9) | 33 (5.1) | 30 (5.7) |
| **Treatment Type**  Adjuvant  Consolidation  Induction  Maintenance  Neoadjuvant  Palliative  Salvage  **Surgery** | 200 (7.7)  6 (0.2)  23 (0.9)  2 (0.1)  94 (3.8)  2057 (83.6)  77 (3.1) | 67 (5.2)  4 (0.3)  5 (0.4)  1 (0.1)  42 (3.3)  1,138 (88.3)  32 (2.5) | 72 (11.1)  1 (0.2)  9 (1.4)  0 (0.0)  29 (4.5)  508 (78.4)  29 (4.5) | 61 (11.7)  1 (0.2)  9 (1.7)  1 (0.2)  23 (4.4)  411 (78.7)  16 (3.1) |
| No | 1,892 (76.9) | 1,048 (81.3) | 483 (74.5) | 361 (69.2) |
| Yes | 567 (23.1) | 241 (18.7) | 165 (25.5) | 161 (30.8) |
| **Radiotherapy** |  |  |  |  |
| No | 1,552 (63.1) | 972 (75.4) | 353 (54.5) | 227 (43.5) |
| Yes | 907 (36.9) | 317 (24.6) | 295 (45.5) | 295 (56.5) |
| Weight | 61.8 ± 10.5 | 60.8 ± 10.4 | 62.9 ± 10.4 | 63.2 ± 10.8 |
| Platelets (10^12 cells/L) | 278.2 ± 112.4 | 282.1 ± 118.8 | 276.1 ± 108.5 | 271.0 ± 100.3 |
| RBC (10^12 cells/L) | 4.4 ± 0.5 | 4.3 ± 0.6 | 4.4 ± 0.5 | 4.5 ± 0.5 |
| WBC (10^3 cells/ul)  Neutrophil (10^3 cells/ul) | 8.4 ± 6.5  5.5 ± 3.3 | 9.1 ± 6.5  5.8 ± 3.6 | 7.9 ± 5.3  5.4 ± 3 | 7.4 ± 7.8  5.0 ± 2.6 |
| Albumin (g/dL) | 3.6 ± 0.7 | 3.5 ± 0.7 | 3.7 ± 0.6 | 3.9 ± 0.6 |
| BSA | 1.7 ± 0.2 | 1.7 ± 0.2 | 1.7 ± 0.2 | 1.7 ± 0.2 |
| Chemo duration | 422.4 ± 544.9 | 153.5 ± 232.1 | 461.2 ± 426.3 | 1,038.2 ± 693.7 |
| Total treatment dosage (mg or mg/m^2^) | 22,008.2 ± 74,070.6 | 9,640.3 ± 26,020.2 | 19,093.0 ± 49,114.8 | 56,167.7 ± 140,202.4 |
| Anti-cancer drug prescription frequency | 21.4 ± 24.1 | 7.3 ± 4.3 | 21.9 ± 4.2 | 55.5 ± 31.7 |
| **Formulation type** |  |  |  |  |
| Injection users | 1,642 (66.8) | 930 (72.1) | 444 (68.5) | 268 (51.3) |
| Oral users | 234 (9.5) | 196 (15.2) | 25 (3.9) | 13 (2.5) |
| Oral & Injection users | 583 (23.7) | 163 (12.6) | 179 (27.6) | 241 (46.2) |
| **Metastatic site Change** |  |  |  |  |
| Nil or no new metastasis | 526 (21.4) | 301 (23.4) | 125 (19.3) | 100 (19.2) |
| 1 | 627 (25.5) | 345 (26.8) | 157 (24.2) | 125 (23.9) |
| 2 | 498 (20.3) | 263 (20.4) | 129 (19.9) | 106 (20.3) |
| 3 | 396 (16.1) | 192 (14.9) | 119 (18.4) | 85 (16.3) |
| 4 | 226 (9.2) | 113 (8.8) | 59 (9.1) | 54 (10.3) |
| 5 | 105 (4.3) | 46 (3.6) | 31 (4.8) | 28 (5.4) |
| 6 | 56 (2.3) | 21 (1.6) | 22 (3.4) | 13 (2.5) |
| 7 | 19 (0.8) | 7 (0.5) | 4 (0.6) | 8 (1.5) |
| 8 | 4 (0.2) | 0 (0.0) | 2 (0.3) | 2 (0.4) |
| 9 | 2 (0.1) | 1 (0.1) | 0 (0.0) | 1 (0.2) |
| **During chemotherapy metastasis** |  |  |  |  |
| No | 1,758 (71.5) | 1,030 (79.9) | 433 (66.8) | 295 (56.5) |
| Yes | 701 (28.5) | 259 (20.1) | 215 (33.2) | 227 (43.5) |
| Alkylating |  |  |  |  |
| No | 624 (25.4) | 496 (38.5) | 75 (11.6) | 53 (10.2) |
| Yes | 1,835 (74.6) | 793 (61.5) | 573 (88.4) | 469 (89.8) |
| Antiandrogen |  |  |  |  |
| No | 2,458 (100.0) | 1,289 (100.0) | 648 (100.0) | 521 (99.8) |
| Yes | 1 (0.0) | 0 (0.0) | 0 (0.0) | 1 (0.2) |
| Antimetabolite |  |  |  |  |
| No | 1,041 (42.3) | 744 (57.7) | 204 (31.5) | 93 (17.8) |
| Yes | 1,418 (57.7) | 545 (42.3) | 444 (68.5) | 429 (82.2) |
| Anti-mitotic |  |  |  |  |
| No | 2,080 (84.6) | 1,202 (93.3) | 517 (79.8) | 361 (69.2) |
| Yes | 379 (15.4) | 87 (6.7) | 131 (20.2) | 161 (30.8) |
| Biologics |  |  |  |  |
| No | 1,924 (78.2) | 1,135 (88.1) | 480 (74.1) | 309 (59.2) |
| Yes | 535 (21.8) | 154 (11.9) | 168 (25.9) | 213 (40.8) |
| Immunosuppressant |  |  |  |  |
| No | 2,439 (99.2) | 1,284 (99.6) | 644 (99.4) | 511 (97.9) |
| Yes | 20 (0.8) | 5 (0.4) | 4 (0.6) | 11 (2.1) |
| Kinase inhibitor |  |  |  |  |
| No | 2,449 (99.6) | 1,288 (99.9) | 645 (99.5) | 516 (98.9) |
| Yes | 10 (0.4) | 1 (0.1) | 3 (0.5) | 6 (1.1) |
| Pyrimidines |  |  |  |  |
| No | 1,855 (75.4) | 1,165 (90.4) | 460 (71.0) | 230 (44.1) |
| Yes | 604 (24.6) | 124 (9.6) | 188 (29.0) | 292 (55.9) |
| Receptor tyrosine kinase inhibitors |  |  |  |  |
| No | 1,667 (67.8) | 935 (72.5) | 449 (69.3) | 283 (54.2) |
| Yes | 792 (32.2) | 354 (27.5) | 199 (30.7) | 239 (45.8) |
| Taxane |  |  |  |  |
| No | 1,385 (56.3) | 796 (61.8) | 353 (54.5) | 236 (45.2) |
| Yes | 1,074 (43.7) | 493 (38.2) | 295 (45.5) | 286 (54.8) |
| Topoisomerase inhibitors |  |  |  |  |
| No | 2,248 (91.4) | 1,235 (95.8) | 587 (90.6) | 426 (81.6) |
| Yes | 211 (8.6) | 54 (4.2) | 61 (9.4) | 96 (18.4) |
| **Chemo Class** |  |  |  |  |
| Double chemo classes | 822 (33.4) | 623 (48.3) | 164 (25.3) | 35 (6.7) |
| Quadruple chemo classes | 335 (13.6) | 40 (3.1) | 149 (23.0) | 146 (28.0) |
| Quintuple or above classes | 234 (9.5) | 6 (0.5) | 43 (6.6) | 185 (35.4) |
| Single chemo class | 445 (18.1) | 394 (30.6) | 34 (5.2) | 17 (3.3) |
| Triple chemo classes | 623 (25.3) | 226 (17.5) | 258 (39.8) | 139 (26.6) |
| Comorbidity |  |  |  |  |
| No | 1,323 (53.8) | 747 (58.0) | 338 (52.2) | 238 (45.6) |
| Yes | 1,136 (46.2) | 542 (42.0) | 310 (47.8) | 284 (54.4) |
| Diabetes |  |  |  |  |
| No | 2,063 (83.9) | 1,107 (85.9) | 535 (82.6) | 421 (80.7) |
| Yes | 396 (16.1) | 182 (14.1) | 113 (17.4) | 101 (19.3) |
| Anemia |  |  |  |  |
| No | 2,293 (93.2) | 1,220 (94.6) | 608 (93.8) | 465 (89.1) |
| Yes | 166 (6.8) | 69 (5.4) | 40 (6.2) | 57 (10.9) |
| Fatty liver |  |  |  |  |
| No | 2,438 (99.1) | 1,279 (99.2) | 641 (98.9) | 518 (99.2) |
| Yes | 21 (0.9) | 10 (0.8) | 7 (1.1) | 4 (0.8) |
| Chronic liver disease |  |  |  |  |
| No | 2,402 (97.7) | 1,269 (98.4) | 635 (98.0) | 498 (95.4) |
| Yes | 57 (2.3) | 20 (1.6) | 13 (2.0) | 24 (4.6) |
| Anxiety |  |  |  |  |
| No | 2,266 (92.2) | 1,202 (93.3) | 590 (91.0) | 474 (90.8) |
| Yes | 193 (7.8) | 87 (6.7) | 58 (9.0) | 48 (9.2) |
| Obsessive compulsive disorder |  |  |  |  |
| No | 2,457 (99.9) | 1,287 (99.8) | 648 (100.0) | 522 (100.0) |
| Yes | 2 (0.1) | 2 (0.2) | 0 (0.0) | 0 (0.0) |
| Panic disorder |  |  |  |  |
| No | 2,456 (99.9) | 1,288 (99.9) | 647 (99.8) | 521 (99.8) |
| Yes | 3 (0.1) | 1 (0.1) | 1 (0.2) | 1 (0.2) |
| Chronic obstructive pulmonary disease |  |  |  |  |
| No | 2,076 (84.4) | 1,084 (84.1) | 542 (83.6) | 450 (86.2) |
| Yes | 383 (15.6) | 205 (15.9) | 106 (16.4) | 72 (13.8) |
| Chronic kidney disease |  |  |  |  |
| No | 2,392 (97.3) | 1,263 (98.0) | 630 (97.2) | 499 (95.6) |
| Yes | 67 (2.7) | 26 (2.0) | 18 (2.8) | 23 (4.4) |
| Renal failure |  |  |  |  |
| No | 2,344 (95.3) | 1,237 (96.0) | 619 (95.5) | 488 (93.5) |
| Yes | 115 (4.7) | 52 (4.0) | 29 (4.5) | 34 (6.5) |
| Coronary artery disease |  |  |  |  |
| No | 2,294 (93.3) | 1,209 (93.8) | 603 (93.1) | 482 (92.3) |
| Yes | 165 (6.7) | 80 (6.2) | 45 (6.9) | 40 (7.7) |
| Hypothyroidism |  |  |  |  |
| No | 2,331 (94.8) | 1,245 (96.6) | 611 (94.3) | 475 (91.0) |
| Yes | 128 (5.2) | 44 (3.4) | 37 (5.7) | 47 (9.0) |
| Hyperthyroidism |  |  |  |  |
| No | 2,429 (98.8) | 1,276 (99.0) | 637 (98.3) | 516 (98.9) |
| Yes | 30 (1.2) | 13 (1.0) | 11 (1.7) | 6 (1.1) |
| Lobectomy (liver) |  |  |  |  |
| No | 2,193 (89.2) | 1,172 (90.9) | 568 (87.7) | 453 (86.8) |
| Yes | 266 (10.8) | 117 (9.1) | 80 (12.3) | 69 (13.2) |
| Lung transplant |  |  |  |  |
| No | 2,458 (100.0) | 1,288 (99.9) | 648 (100.0) | 522 (100.0) |
| Yes | 1 (0.0) | 1 (0.1) | 0 (0.0) | 0 (0.0) |
| Lymph node dissection |  |  |  |  |
| No | 2,142 (87.1) | 1,151 (89.3) | 553 (85.3) | 438 (83.9) |
| Yes | 317 (12.9) | 138 (10.7) | 95 (14.7) | 84 (16.1) |
| Pleurectomy |  |  |  |  |
| No | 2,458 (100.0) | 1,288 (99.9) | 648 (100.0) | 522 (100.0) |
| Yes | 1 (0.0) | 1 (0.1) | 0 (0.0) | 0 (0.0) |
| Pneumonectomy |  |  |  |  |
| No | 2,435 (99.0) | 1,279 (99.2) | 642 (99.1) | 514 (98.5) |
| Yes | 24 (1.0) | 10 (0.8) | 6 (0.9) | 8 (1.5) |
| Segmentectomy |  |  |  |  |
| No | 2,451 (99.7) | 1,284 (99.6) | 646 (99.7) | 521 (99.8) |
| Yes | 8 (0.3) | 5 (0.4) | 2 (0.3) | 1 (0.2) |
| Thoracotomy |  |  |  |  |
| No | 2,452 (99.7) | 1,286 (99.8) | 645 (99.5) | 521 (99.8) |
| Yes | 7 (0.3) | 3 (0.2) | 3 (0.5) | 1 (0.2) |
| Thymectomy |  |  |  |  |
| No | 2,458 (100.0) | 1,288 (99.9) | 648 (100.0) | 522 (100.0) |
| Yes | 1 (0.0) | 1 (0.1) | 0 (0.0) | 0 (0.0) |
| Other procedure |  |  |  |  |
| No | 2,099 (85.4) | 1,132 (87.8) | 552 (85.2) | 415 (79.5) |
| Yes | 360 (14.6) | 157 (12.2) | 96 (14.8) | 107 (20.5) |
| Wedge resection |  |  |  |  |
| No | 2,320 (94.3) | 1,223 (94.9) | 613 (94.6) | 484 (92.7) |
| Yes | 139 (5.7) | 66 (5.1) | 35 (5.4) | 38 (7.3) |
| ^1^n (%); Mean ± SD | | | | |

**Supplementary table 2:** Post-stratification non-small cell lung cancer patient (N=2459) characteristics.

| **Characteristic** | **Overall**  N = 445^1^ | **Group-B**  N = 34^1^ | **Group-A**  N = 394^1^ | **Group-C** N = 17^1^ |
| --- | --- | --- | --- | --- |
| First Line Singlet Type |  |  |  |  |
| Alkylating | 15 (3.4) | 0 (0.0) | 15 (3.8) | 0 (0.0) |
| Anti-mitotic | 9 (2.0) | 1 (2.9) | 8 (2.0) | 0 (0.0) |
| Antimetabolite | 78 (17.5) | 4 (11.8) | 71 (18.0) | 3 (17.6) |
| Biologics | 42 (9.4) | 2 (5.9) | 39 (9.9) | 1 (5.9) |
| Pyrimidines | 22 (4.9) | 2 (5.9) | 20 (5.1) | 0 (0.0) |
| Taxane | 47 (10.6) | 1 (2.9) | 45 (11.4) | 1 (5.9) |
| Topoisomerase I inhibitor | 4 (0.9) | 0 (0.0) | 4 (1.0) | 0 (0.0) |
| Topoisomerase II inhibitor | 1 (0.2) | 0 (0.0) | 1 (0.3) | 0 (0.0) |
| Receptor tyrosine kinase inhibitor | 227 (51.0) | 24 (70.6) | 191 (48.5) | 12 (70.6) |
| Post chemotherapy OS | 12.9 ± 18.2 | 31.0 ± 19.6 | 9.1 ± 11.4 | 64.9 ± 36.7 |
| ^1^n (%); Mean ± SD | | | | |

**Supplementary table 3:** List of singlet chemo classes applied on NSCLC (n=445) patients. All these patients received single chemo class until death.

| **Characteristic** | **Overall**  N = 822^1^ | **Group-B**  N = 164^1^ | **Group-A**  N = 623^1^ | **Group-C**  N = 35^1^ |
| --- | --- | --- | --- | --- |
| First Line Doublet User |  |  |  |  |
| Alkylating, Anti-mitotic | 53 (6.4) | 14 (8.5) | 39 (6.3) | 0 (0.0) |
| Alkylating, Pyrimidines | 77 (9.4) | 26 (15.9) | 46 (7.4) | 5 (14.3) |
| Alkylating, Taxane | 288 (35.0) | 31 (18.9) | 255 (40.9) | 2 (5.7) |
| Antimetabolite, Alkylating | 201 (24.5) | 38 (23.2) | 152 (24.4) | 11 (31.4) |
| Antimetabolite, Taxane | 20 (2.4) | 2 (1.2) | 18 (2.9) | 0 (0.0) |
| Antimetabolite, Receptor tyrosine kinase inhibitor | 27 (3.3) | 3 (1.8) | 21 (3.4) | 3 (8.6) |
| Other Combinations | 82 (10.0) | 15 (9.1) | 58 (9.3) | 9 (25.7) |
| Topoisomerase II Inhibitor, Alkylating | 74 (9.0) | 35 (21.3) | 34 (5.5) | 5 (14.3) |
| Post chemotherapy OS | 10.9 ± 14.8 | 16.2 ± 17.6 | 8.3 ± 12.1 | 32.0 ± 20.3 |
| ^1^n (%); Mean ± SD | | | | |

**Supplementary table 4:** List of various doublet agents applied on NSCLC (n=822) patients. All these patients received double chemo classes until death.

| **Characteristic** | **Overall**  N = 623^1^ | **Group-B**  N = 258^1^ | **Group-A**  N = 226^1^ | **Group-C**  N = 139^1^ |
| --- | --- | --- | --- | --- |
| First Line combinations of Triplet User |  |  |  |  |
| Alkylating, Anti-mitotic | 46 (7.4) | 29 (11.2) | 6 (2.7) | 11 (7.9) |
| Alkylating, Pyrimidines | 42 (6.7) | 22 (8.5) | 8 (3.5) | 12 (8.6) |
| Alkylating, Taxane | 215 (34.5) | 90 (34.9) | 92 (40.7) | 33 (23.7) |
| Antimetabolite, Alkylating | 143 (23.0) | 62 (24.0) | 61 (27.0) | 20 (14.4) |
| Antimetabolite, Taxane | 5 (0.8) | 1 (0.4) | 1 (0.4) | 3 (2.2) |
| Other Doublet Combination | 101 (16.2) | 29 (11.2) | 42 (18.6) | 30 (21.6) |
| Topoisomerase II Inhibitor, Alkylating | 34 (5.5) | 12 (4.7) | 2 (0.9) | 20 (14.4) |
| Receptor tyrosine kinase inhibitor, Antimetabolite | 37 (5.9) | 13 (5.0) | 14 (6.2) | 10 (7.2) |
| Triplet Users Triplet Type |  |  |  |  |
| Other triplets | 208 (33.4) | 83 (32.2) | 83 (36.7) | 42 (30.2) |
| Triplet as Biologics | 125 (20.1) | 56 (21.7) | 49 (21.7) | 20 (14.4) |
| Triplet as Pyrimidines | 72 (11.6) | 36 (14.0) | 10 (4.4) | 26 (18.7) |
| Triplet as RTKI | 121 (19.4) | 44 (17.1) | 57 (25.2) | 20 (14.4) |
| Triplet as Taxane | 64 (10.3) | 29 (11.2) | 22 (9.7) | 13 (9.4) |
| Triplet as Topoisomerase I/II inhibitors | 33 (5.3) | 10 (3.9) | 5 (2.2) | 18 (12.9) |
| Post chemotherapy OS | 19.3 ± 18.9 | 18.3 ± 14.2 | 10.7 ± 14.7 | 35.0 ± 22.8 |
| ^1^n (%); Mean ± SD | | | | |

**Supplementary table 5:** List of various triplet agents applied on NSCLC (n=623) patients. All these patients received doublet & triplet chemo classes until death. Tyrosine kinase inhibitors are same as (RTKI) mentioned in abbreviation section of main text.

| **Characteristic** | **Overall**  N = 569^1^ | **Group-B**  N = 192^1^ | **Group-A**  N = 46^1^ | **Group-C**  N = 331^1^ |
| --- | --- | --- | --- | --- |
| Second Line Users First Line Doublet Type |  |  |  |  |
| Alkylating, Anti-mitotic | 61 (10.7) | 21 (10.9) | 1 (2.2) | 39 (11.8) |
| Alkylating, Antimetabolite | 126 (22.1) | 50 (26.0) | 8 (17.4) | 68 (20.5) |
| Alkylating, Taxane | 167 (29.3) | 70 (36.5) | 12 (26.1) | 85 (25.7) |
| Alkylating, Topoisomerase II Inhibitor | 13 (2.3) | 5 (2.6) | 0 (0.0) | 8 (2.4) |
| Antimetabolite, Taxane | 9 (1.6) | 1 (0.5) | 0 (0.0) | 8 (2.4) |
| Other combinations as first line doublet | 61 (10.7) | 14 (7.3) | 10 (21.7) | 37 (11.2) |
| Pyrimidines, Alkylating | 34 (6.0) | 9 (4.7) | 3 (6.5) | 22 (6.6) |
| Receptor tyrosine kinase inhibitor, Alkylating | 49 (8.6) | 8 (4.2) | 6 (13.0) | 35 (10.6) |
| Receptor tyrosine kinase inhibitor, Antimetabolite | 49 (8.6) | 14 (7.3) | 6 (13.0) | 29 (8.8) |
| Second Line User Quadruplet Type |  |  |  |  |
| Alkylating, Antimetabolite | 11 (1.9) | 4 (2.1) | 1 (2.2) | 6 (1.8) |
| Alkylating, Biologics | 33 (5.8) | 10 (5.2) | 4 (8.7) | 19 (5.7) |
| Alkylating, Pyrimidines | 18 (3.2) | 3 (1.6) | 0 (0.0) | 15 (4.5) |
| Anti-mitotic, Biologics | 33 (5.8) | 17 (8.9) | 1 (2.2) | 15 (4.5) |
| Antimetabolite, Biologics | 44 (7.7) | 11 (5.7) | 5 (10.9) | 28 (8.5) |
| Antimetabolite, Pyrimidines | 38 (6.7) | 14 (7.3) | 5 (10.9) | 19 (5.7) |
| Antimetabolite, Taxane | 18 (3.2) | 7 (3.6) | 2 (4.3) | 9 (2.7) |
| Antimetabolite, Receptor tyrosine kinase inhibitor | 82 (14.4) | 34 (17.7) | 7 (15.2) | 41 (12.4) |
| Biologics, Pyrimidines | 54 (9.5) | 19 (9.9) | 0 (0.0) | 35 (10.6) |
| Other Combination as quadruplet | 56 (9.9) | 12 (6.3) | 3 (6.5) | 41 (12.4) |
| Pyrimidines, Anti-mitotic | 24 (4.2) | 7 (3.6) | 2 (4.3) | 15 (4.5) |
| Pyrimidines, Taxane | 26 (4.6) | 10 (5.2) | 2 (4.3) | 14 (4.2) |
| Taxane, Alkylating | 16 (2.8) | 3 (1.6) | 2 (4.3) | 11 (3.3) |
| Taxane, Anti-mitotic | 13 (2.3) | 2 (1.0) | 1 (2.2) | 10 (3.0) |
| Taxane, Biologics | 43 (7.6) | 19 (9.9) | 4 (8.7) | 20 (6.0) |
| Taxane, Tyrosine kinase inhibitor | 20 (3.5) | 4 (2.1) | 2 (4.3) | 14 (4.2) |
| Receptor tyrosine kinase inhibitor, Biologics | 18 (3.2) | 8 (4.2) | 4 (8.7) | 6 (1.8) |
| Receptor tyrosine kinase inhibitor, Pyrimidines | 22 (3.9) | 8 (4.2) | 1 (2.2) | 13 (3.9) |
| Post chemotherapy OS | 29.1 ± 21.7 | 20.7 ± 16.9 | 11.4 ± 11.1 | 36.4 ± 22.2 |
| ^1^n (%); Mean ± SD | | | | |

**Supplementary table 6:** List of various quadruplet agents applied on NSCLC (n=569) patients. All these patients received doublet & quadruplet chemo classes until death. Tyrosine kinase inhibitors are same as (RTKI) mentioned in abbreviation section of main text.

| **Year wise Deaths** | **Overall**  **N = 2459^1^** | **Adjuvant**  **N = 200^1^** | **Consolidation**  **N = 6^1^** | **Induction**  **N = 23^1^** | **Maintenance**  **N = 2^1^** | **Neoadjuvant**  **N - 94^1^** | **Palliative**  **N = 2057^1^** | **Salvage**  **N = 77^1^** |
| --- | --- | --- | --- | --- | --- | --- | --- | --- |
| Group-A  1-Year Deaths | N = 1,289  1,002 (77.7) | N = 67  36 (53.7) | N = 4  3 (75.0) | N = 5  4 (80.0) | N = 1  0 (0.0) | N = 42  31 (73.8) | N = 1,138  905 (79.5) | N = 32  23 (71.9) |
| 2-Year Deaths | 190 (14.7) | 10 (14.9) | 0 (0.0) | 1 (20.0) | 0 (0.0) | 9 (21.4) | 162 (14.2) | 8 (25.0) |
| 3-Year Deaths | 51 (4.0) | 8 (11.9) | 0 (0.0) | 0 (0.0) | 1 (100.0) | 1 (2.4) | 41 (3.6) | 0 (0.0) |
| 4-Year Deaths | 24 (1.9) | 3 (4.5) | 1 (25.0) | 0 (0.0) | 0 (0.0) | 0 (0.0) | 19 (1.7) | 1 (3.1) |
| 5-Year Deaths  >6 Year Deaths  Group-B  1-Year Deaths  2-Year Deaths  3-Year Deaths  4-Year Deaths  5-Year Deaths  >6 Year Deaths  Group-C  1-Year Deaths  2-Year Deaths  3-Year Deaths  4-Year Deaths  5-Year Deaths  >6 Year Deaths | 7 (0.5)  15 (1.2)  N = 648  294 (45.4)  183 (28.2)  89 (13.7)  44 (6.8)  18 (2.8)  20 (3.1)  N = 522  48 (9.2)  142 (27.2)  126 (24.1)  87 (16.7)  36 (6.9)  83 (15.9) | 2 (3.0)  8 (11.9)  N = 72  18 (25.0)  23 (31.9)  12 (16.7)  9 (12.5)  4 (5.6)  6 (8.3)  N = 61  2 (3.3)  10 (16.4)  16 (26.2)  15 (24.6)  3 (4.9)  15 (24.6) | 0 (0.0)  0 (0.0)  N = 1  1 (100.0)  0 (0.0)  0 (0.0)  0 (0.0)  0 (0.0)  0 (0.0)  N = 1  0 (0.0)  0 (0.0)  1 (100.0)  0 (0.0)  0 (0.0)  0 (0.0) | 0 (0.0)  0 (0.0)  N = 9  3 (33.3)  4 (44.4)  1 (11.1)  1 (11.1)  0 (0.0)  0 (0.0)  N = 9  0 (0.0)  2 (22.2)  2 (22.2)  1 (11.1)  0 (0.0)  4 (44.4) | 0 (0.0)  0 (0.0)  N = 0  0 (0.0)  0 (0.0)  0 (0.0)  0 (0.0)  0 (0.0)  0 (0.0)  N = 1  0 (0.0)  0 (0.0)  1 (100.0)  0 (0.0)  0 (0.0)  0 (0.0) | 0 (0.0)  1 (2.4)  N = 29  12 (41.4)  8 (27.6)  4 (13.8)  2 (6.9)  1 (3.4)  2 (6.9)  N = 23  2 (8.7)  7 (30.4)  6 (26.1)  5 (21.7)  0 (0.0)  3 (13.0) | 5 (0.4)  6 (0.5)  N = 508  247 (48.6)  136 (26.8)  69 (13.6)  31 (6.1)  13 (2.6)  12 (2.4)  N = 411  42 (10.2)  119 (29.0)  96 (23.4)  64 (15.6)  31 (7.5)  59 (14.4) | 0 (0.0)  0 (0.0)  N = 29  13 (44.8)  12 (41.4)  13 (44.8)  1 (3.4)  0 (0.0)  0 (0.0)  N = 16  2 (12.5)  4 (25.0)  4 (25.0)  2 (12.5)  2 (12.5)  2 (12.5) |
| ^1^n (%) | | | | | | | | |

**Supplementary table 7: Year wise deaths as per cancer treatment type received by NSCLC (n=2459) patients.**

| Generic Names | Chemo Class | Formulation |
| --- | --- | --- |
| Etoposide | Topoisomerase II Inhibitor | Injection |
| Cisplatin | Alkylating | Injection |
| Pemetrexed | Antimetabolite | Injection |
| Nivolumab | Biologics | Injection |
| Carboplatin | Alkylating | Injection |
| Paclitaxel | Taxane | Injection |
| Atezolizumab | Biologics | Injection |
| Gemcitabine | Pyrimidines | Injection |
| Vinorelbine | Anti-mitotic | Injection |
| Docetaxel | Taxane | Injection |
| Pembrolizumab | Biologics | Injection |
| Gefitinib | Tyrosine kinase inhibitor | Oral |
| Osimertinib | Tyrosine kinase inhibitor | Oral |
| Topotecan | Topoisomerase I Inhibitor | Oral |
| Crizotinib | Tyrosine kinase inhibitor | Oral |
| Afatinib | Tyrosine kinase inhibitor | Oral |
| Irinotecan | Topoisomerase I Inhibitor | Injection |
| Trastuzumab | Biologics | Injection |
| Capecitabine | Pyrimidines | Oral |
| fluorouracil | Pyrimidines | Injection |
| Erlotinib | Tyrosine kinase inhibitor | Oral |
| Ifosfamide | Alkylating | Injection |
| Iretinib | Tyrosine kinase inhibitor | Oral |
| Ceritinib | Tyrosine kinase inhibitor | Oral |
| Bevacizumab | Biologics | Injection |
| Doxorubicin | Topoisomerase II Inhibitor | Injection |
| Vincristine | Anti-mitotic | Injection |
| Cyclophosphamide | Immunosuppressant | Injection |
| Topotecan | Topoisomerase I Inhibitor | Injection |
| Sunitinib | Tyrosine kinase inhibitor | Oral |
| Everolimus | Immunosuppressant | Oral |
| Gimeracil, oteracil, Tegafur | Pyrimidines | Oral |
| Oxaliplatin | Alkylating | Injection |
| Belotecan | Topoisomerase I Inhibitor | Injection |
| Durvalumab | Biologics | Injection |
| Ramucirumab | Biologics | Injection |
| Alectinib | Tyrosine kinase inhibitor | Oral |
| Dabrafenib | Kinase inhibitor | Oral |
| Trametinib | Kinase inhibitor | Oral |
| Ipilimumab | Biologics | Injection |
| Eribulin | Anti-mitotic | Injection |
| Cytarabine | Pyrimidines | Injection |
| Enzalutamide | Antiandrogen | Oral |
| Olaratumab | Biologics | Injection |
| Lazertinib | Tyrosine kinase inhibitor | Oral |
| Cetuximab | Biologics | Injection |
| Lenvatinib | Tyrosine kinase inhibitor | Oral |
| Pazopanib | Tyrosine kinase inhibitor | Oral |
| Rituximab | Biologics | Injection |
| Imatinib | Tyrosine kinase inhibitor | Oral |
| Azacitidine | Antimetabolite | Injection |
| Decitabine | Pyrimidines | Injection |
| Axitinib | Tyrosine kinase inhibitor | Oral |
| Lorlatinib | Kinase inhibitor | Oral |
| Methotrexate | Immunosuppressant | Injection |
| Vinblastine | Anti-mitotic | Injection |
| Lubinectedin | Alkylating | Injection |
| Brigatinib | Tyrosine kinase inhibitor | Oral |

**Supplementary table 8:** List of generic names, chemo-class, and formulation type of drugs prescribed to NSCLC patients. Using generic names anti-cancer classes were derived and formulation type was derived from trade names available in EMR. There are more than 195 trade names with different formulation type of above mentioned 58 generic products.

**Other Surgery Types**

Excision of chest wall mass by thoracotomy

VATS ()

Empyemectomy under VATS

Pleural biopsy under VATS

Open lung biopsy

Incisional biopsy of mediastinum

Mediastinal mass excision under VATS

Bullectomy under VATS, left

Mediastinoscopy

Excision of mediastinal mass, right

Pleural biopsy

Mediastinal dissection

Rib resection under VATS (video assisted thoracoscopic surgery)

VATS (video assisted thoracoscopic surgery)-Bullectomy, right

Excision of lesion of mediastinum

Mediastinal tumor excision under VATS

Lung biopsy

Excision of mediastinal tumor

Cervical mediastinoscopy

Wedge biopsy of liver

Thoracoscopic pleurodesis

Lung biopsy under VATS

Thoracoscopic pleural biopsy

Bullectomy, right

Excision of chest wall mass(malignant)(soft tissue)

Pleurolysis, left

Repair of diaphragm, right

Biopsy by mediastinoscopy

Mediastinal mass excision under VATS, right

Subtotal esophagectomy under VATS (video assisted thoracoscopic surgery)

pleural mass excision

Thoracoscopic wedge biopsy of lung

Mediastinotomic biopsy

Pleural mass excision under VATS (video assisted thoracoscopic surgery)

Excision of mediastinal malignant tumor

Mass excision, chest wall

Incision and Drainage

Repair of diaphragmatic hernia, left

Excision of mediastinal cyst

Biopsy of mediastinum by endoscope

Bleeder ligation (heart and lung postop)

Empyema cavity irrigation under VATS

Pulmonary arterigraphy

Excision of chest wall mass(benign)

Mediastinal tumor excision

VATS (video assisted thoracoscopic surgery)-Bullectomy

Mediastinotomy

Partial resection, rib, left

Decortication under VATS

Pleurolysis, right

Mediastinal mass excision under VATS, left

(SP) Robot assisted thoracoscopic excision of mediastinal mass (complex) (gradeⅠ)((SP)

Open pleural biopsy

Bullectomy under VATS

Excision of pleural mass

Rib resection (2nd rib)

Empyemectomy under VATS, left

Pleurodesis

Excision of mediastinal mass, left

Decortication of lung, left

Thoracoscopy

Broncho-pleural fistula closure (Repair of BPF)

Excision of mediastinal tumor under VATS

Chest tube insertion, bilateral

Rib resection

Partial rib resection

Pleural biopsy by explo thoracotomy

Enucleation of lung mass

Postopertive bleeder ligation (heart and lung postop)

Decortication under VATS, left

Empyemectomy under VATS, right

Incisional biopsy of chest wall mass

Irrigation and Drainage

Percutaneous cardiopulmonary bypass

VATS (video assisted thoracoscopic surgery)-Bullectomy, left

Pleural mass excision under VATS (video assisted thoracoscopic surgery), left

Excision of bronchus, wide sleeve

Pulmonary artery thrombectomy

Lung abscess incision

Suture of lung

Needle biopsy of lung

Open reduction of rib fracture

Removal of chest wall foreign body(other)

Open lung biopsy under VATS

Mediastinal tumor excision, Rt

Mediastinal tracheostomy

Decortication of lung

Decortication of lung and parietal pleurectomy

Mediastinoscopic lung biopsy

Sympathicotomy under VATS

En-bloc resection of chest wall(left)

Diaphragm repair under VATS

Partial resection of mediastinal tumor

Excision of mediastinal mass, left

Decortication under VATS, right

Acellular Dermal Graft [other, over 100cm2~under 400cm2]

Repair of diaphragmatic hernia

Chest tube insertion

Repair of diaphragm, left

Excision of bronchial tumor

Lung biopsy, open

VATS (video assisted thoracoscopic surgery)-Excision of lesion of diaphragm

Pleurolysis by VATS (video assisted thoracoscopic surgery)

Bullectomy, left

Mediastinal bleeder ligation

Other diagnostic procedures on mediastinum

Thoracoscopic lung biopsy

Bullectomy

Acellular Dermal Graft [other, under 25cm2]

Abscess incision and Drainage

Chest wall mass excision under VATS (video assisted thoracoscopic surgery), right

Partial resection of the rib

Decortication of lung, right

Excision of chest wall

Incision and drainage of chest wall

Excision of bulla of lung

Mediastinal mass excision via lateral thoracotomy

Chest wall mass excision (soft tissue malignant tumor)

Segmental rib resection

Excision of chest wall mass(malignant) by thoracotomy

Rib resection(other)

Repair of diaphragmatic hernia (left)

Mediastinotomy for drainage(thoracotomy)

Diaphragmatic plication under VATS

Mediastinotomy for drainage(closed)

Open lung biopsy by thoracotomy

Anterior mediastinostomy by thoracotomy

Rib biopsy,open

Syringo-pleural shunt

Repair of direct inguinal hernia, left

Pleural mass excision, right

Puncture bx. of kidney, liver, lung

VATS (video assisted thoracoscopic surgery)-Incisional biopsy of mediastinum

Repair of diaphragmatic hernia, abdominal approach, left

Lung suture

Robot assisted thoracoscopic excision of mediastinal mass (simple) (grade I)

Pulmonary artery thromboendarterectomy

Excision of mediastinal mass,left

Repair of direct inguinal hernia, right

mediastinotomy for drainage

Radiofrequency ablation of lung

Bullectomy under VATS, right

Photodynamic therapy under VATS

Deep vein thrombectomy(chest)

Mediastinotomy for drainage

Chest wall reconstruction, right

T3 sympathicotomy under VATS, bilateral

VATS (video assisted thoracoscopic surgery)-Pericardial window formation

Pulmonary valvulotomy

Robot assisted thoracoscopic excision of mediastinal mass (complex) (grade I)

Pulmonary arterioplasty

Robot assisted thoracoscopic excision of mediastinal mass (simple) (benign)

Robot assisted thoracoscopic excision of mediastinal mass (grade II)

Sympathectomy under VATS (video assisted thoracoscopic surgery)

Chest wall mass excision under VATS (video assisted thoracoscopic surgery)

Enucleation of hamartoma, lung

Chest wall mass excision under VATS (video assisted thoracoscopic surgery), left

Single-port thoracoscopic pleural biopsy, left

**Supplementary table 9:** List of 158 surgeries considered as other procedures.
