## Supplementary File 2 for "Prognostic Features of Anti-Cancer Drugs Response in Resected/Unresected Primary Non-Small Cell Lung Cancer: A Retrospective Cohort Study"

Saikat Samadder^1*^

^1^Department of Pharmacology, Yonsei University College of Medicine, 50 Yonsei-ro, Seodaemun-Gu, Seoul, Korea

^*^ Correspondence: Saikat Samadder

**Supplementary file 2**

**Table of contents**

**1. Supplementary** **Table 1: Year wise mortality in Group-A singlet chemo class users as first-line.**

**Supplementary** **Table 2: Year wise mortality in Group-B singlet chemo class users as first-line.**

**Supplementary Table 3: Year wise mortality in Group-C singlet chemo class users as first-line.**

**Supplementary Table 3: Year wise mortality in Group-C singlet chemo class users as first-line.**

**Supplementary** **Table 4: Year wise mortality in Group-A first-line doublet chemo class users. ------------------------------------------------------------------------------------------------------------------------------- Page [2]**

**2. Supplementary** **Table 5: Year wise mortality in Group-B first-line doublet chemo class users as first-line.**

**Supplementary** **Table 6: Year wise mortality in Group-C first-line Doublet chemo class users as first-line.**

**Supplementary** **Table 7: Year wise mortality in Group-A First-Line Doublet combination of triplet users (Table7A) and overall triplet chemo class type (Table7B). --------------------------------------- Page [3]**

**3. Supplementary** **Table 8: Year wise mortality in Group-C First-Line Doublet chemo class of triplet users (Table8A) and overall triplet chemo class type (Table8B).**

**Supplementary** **Table 9: Year wise mortality in Group-C First-Line Doublet chemo class of triplet users (Table9A) and overall triplet chemo class** type **(Table9B). ---------------------------------------- Page [4]**

**3. Supplementary** **Table 10: Year wise mortality in Group-A quadruplet users. First-line chemo of quadruplet chemo class users.**

**Supplementary** **Table 11: Year wise mortality in Group-A quadruplet users. Second-line chemo of quadruplet chemo class users.**

**Supplementary** **Table 12: Year wise mortality in Group-B quadruplet users. First-line chemo of quadruplet chemo class users. ------------------------------------------------------------------------------------ Page [5]**

**4. Supplementary** **Table 13: Year wise mortality in Group-B quadruplet users. Second-line chemo of quadruplet chemo class users.**

**Supplementary** **Table 14: Year wise mortality in Group-C quadruplet users. First-line chemo of quadruplet chemo class users.**

**Supplementary** **Table 15: Year wise mortality in Group-C quadruplet users. Second-line chemo of quadruplet chemo class users. ---------------------------------------------------------------------------------- Page [6]**

**5. Supplementary** **Table 16: Year wise mortality rate in Group-A, B, & C as per comorbidities diagnosed pre/post primary lung cancer** **----------------------------------------------------------------------------** **[Page 7]**

| **Year wise Death** | **Overall**  N = 394^1^ | **Alkylating**  N = 15^1^ | **Anti-mitotic**  N = 8^1^ | **Antimetabolite**  N = 71^1^ | **Biologics**  N = 39^1^ | **Pyrimidines**  N = 20^1^ | **Taxane**  N = 45^1^ | **Topoisomerase I Inhibitor**  N = 4^1^ | **Topoisomerase II Inhibitor**  N = 1^1^ | **Tyrosine kinase inhibitor**  N = 191^1^ |
| --- | --- | --- | --- | --- | --- | --- | --- | --- | --- | --- |
| Group-A OS |  |  |  |  |  |  |  |  |  |  |
| 1-year death | 293 (74.4) | 15 (100.0) | 8 (100.0) | 58 (81.7) | 36 (92.3) | 16 (80.0) | 38 (84.4) | 4 (100.0) | 1 (100.0) | 117 (61.3) |
| 2-Year death | 69 (17.5) | 0 (0.0) | 0 (0.0) | 10 (14.1) | 2 (5.1) | 2 (10.0) | 6 (13.3) | 0 (0.0) | 0 (0.0) | 49 (25.7) |
| 3-year death | 16 (4.1) | 0 (0.0) | 0 (0.0) | 2 (2.8) | 1 (2.6) | 0 (0.0) | 1 (2.2) | 0 (0.0) | 0 (0.0) | 12 (6.3) |
| 4-Year death | 10 (2.5) | 0 (0.0) | 0 (0.0) | 1 (1.4) | 0 (0.0) | 1 (5.0) | 0 (0.0) | 0 (0.0) | 0 (0.0) | 8 (4.2) |
| 5-Year death  >5-year death | 3 (0.8)  3 (0.8) | 0 (0.0)  0 (0.0) | 0 (0.0)  0 (0.0) | 0 (0.0)  0 (0.0) | 0 (0.0)  0 (0.0) | 0 (0.0)  1 (5.0) | 0 (0.0)  0 (0.0) | 0 (0.0)  0 (0.0) | 0 (0.0)  0 (0.0) | 3 (1.6)  2 (1.0) |
| ^1^n (%) | | | | | | | | | | |

**Supplementary** **Table 1: Year wise mortality in Group-A singlet chemo class users as first-line.**

| **Year wise Death** | **Overall**  N = 34^1^ | **Anti-mitotic**  N = 1^1^ | **Antimetabolite**  N = 4^1^ | **Biologics**  N = 2^1^ | **Pyrimidines**  N = 2^1^ | **Taxane**  N = 1^1^ | **Tyrosine kinase inhibitor**  N = 24^1^ |
| --- | --- | --- | --- | --- | --- | --- | --- |
| Group-B OS |  |  |  |  |  |  |  |
| 1-year death | 5 (14.7) | 0 (0.0) | 0 (0.0) | 1 (50.0) | 0 (0.0) | 1 (100.0) | 3 (12.5) |
| 2-Year death | 9 (26.5) | 1 (100.0) | 4 (100.0) | 0 (0.0) | 1 (50.0) | 0 (0.0) | 3 (12.5) |
| 3-Year death | 7 (20.6) | 0 (0.0) | 0 (0.0) | 0 (0.0) | 0 (0.0) | 0 (0.0) | 7 (29.2) |
| 4-Year death | 8 (23.5) | 0 (0.0) | 0 (0.0) | 0 (0.0) | 0 (0.0) | 0 (0.0) | 8 (33.3) |
| 5-Year death  >5-year death | 3 (8.8)  2 (5.9) | 0 (0.0)  0 (0.0) | 0 (0.0)  0 (0.0) | 0 (0.0)  1 (50.0) | 1 (50.0)  0 (0.0) | 0 (0.0)  0 (0.0) | 2 (8.3)  1 (4.2) |
| ^1^n (%) | | | | | | | |

**Supplementary** **Table 2: Year wise mortality in Group-B singlet chemo class users as first-line.**

| **Year wise Death** | **Overall**  N = 17^1^ | **Antimetabolite**  N = 3^1^ | **Biologics**  N = 1^1^ | **Taxane**  N = 1^1^ | **Tyrosine kinase inhibitor**  N = 12^1^ |
| --- | --- | --- | --- | --- | --- |
| Group-C OS |  |  |  |  |  |
| 1-Year death | 1 (5.9) | 0 (0.0) | 0 (0.0) | 0 (0.0) | 1 (8.3) |
| 2-Year death | 2 (11.8) | 0 (0.0) | 0 (0.0) | 0 (0.0) | 2 (16.7) |
| 3-Year death | 2 (11.8) | 0 (0.0) | 0 (0.0) | 1 (100.0) | 1 (8.3) |
| 4-Year death | 1 (5.9) | 0 (0.0) | 1 (100.0) | 0 (0.0) | 0 (0.0) |
| 5-Year death  >5-year death | 1 (5.9)  10 (58.8) | 0 (0.0)  3 (100.0) | 0 (0.0)  0 (0.0) | 0 (0.0)  0 (0.0) | 1 (8.3)  7 (58.3) |
| ^1^n (%) | | | | | |

**Supplementary Table 3: Year wise mortality in Group-C singlet chemo class users as first-line.**

| **Year wise Death** | **Overall**  N = 623^1^ | **Alkylating, Anti-mitotic**  N = 39^1^ | **Alkylating, Pyrimidines**  N = 46^1^ | **Alkylating, Taxane**  N = 255^1^ | **Antimetabolite, Alkylating**  N = 152^1^ | **Antimetabolite, Taxane**  N = 18^1^ | **Antimetabolite, Tyrosine kinase inhibitor**  N = 21^1^ | **Other Combinations**  N = 58^1^ | **Topoisomerase II Inhibitor, Alkylating**  N = 34^1^ |
| --- | --- | --- | --- | --- | --- | --- | --- | --- | --- |
| Group-A OS |  |  |  |  |  |  |  |  |  |
| 1-Year death | 505 (81.1) | 22 (56.4) | 43 (93.5) | 205 (80.4) | 134 (88.2) | 12 (66.7) | 12 (57.1) | 46 (79.3) | 31 (91.2) |
| 2-Year death | 77 (12.4) | 6 (15.4) | 1 (2.2) | 38 (14.9) | 13 (8.6) | 4 (22.2) | 6 (28.6) | 7 (12.1) | 2 (5.9) |
| 3-Year Death | 23 (3.7) | 6 (15.4) | 1 (2.2) | 4 (1.6) | 4 (2.6) | 1 (5.6) | 3 (14.3) | 4 (6.9) | 0 (0.0) |
| 4-Year Death | 8 (1.3) | 2 (5.1) | 0 (0.0) | 3 (1.2) | 0 (0.0) | 1 (5.6) | 0 (0.0) | 1 (1.7) | 1 (2.9) |
| 5-Year Death  >5-Year Death | 3 (0.5)  7 (1.1) | 2 (5.1)  1 (2.6) | 1 (2.2)  0 (0.0) | 0 (0.0)  5 (2.0) | 0 (0.0)  1 (0.7) | 0 (0.0)  0 (0.0) | 0 (0.0)  0 (0.0) | 0 (0.0)  0 (0.0) | 0 (0.0)  0 (0.0) |
| ^1^n (%) | | | | | | | | | |

**Supplementary** **Table 4: Year wise mortality in Group-A first-line doublet chemo class users.**

| **Year wise Death** | **Overall**  N = 164^1^ | **Alkylating, Anti-mitotic**  N = 14^1^ | **Alkylating, Pyrimidines**  N = 26^1^ | **Alkylating, Taxane**  N = 31^1^ | **Antimetabolite, Alkylating**  N = 38^1^ | **Antimetabolite, Taxane**  N = 2^1^ | **Antimetabolite, Tyrosine kinase inhibitor**  N = 3^1^ | **Other Combinations**  N = 15^1^ | **Topoisomerase II Inhibitor, Alkylating**  N = 35^1^ |
| --- | --- | --- | --- | --- | --- | --- | --- | --- | --- |
| Group-B OS |  |  |  |  |  |  |  |  |  |
| 1-Year Death | 99 (60.4) | 7 (50.0) | 20 (76.9) | 15 (48.4) | 24 (63.2) | 0 (0.0) | 0 (0.0) | 7 (46.7) | 26 (74.3) |
| 2-Year Death | 36 (22.0) | 4 (28.6) | 4 (15.4) | 9 (29.0) | 7 (18.4) | 2 (100.0) | 1 (33.3) | 3 (20.0) | 6 (17.1) |
| 3_Year Death | 15 (9.1) | 1 (7.1) | 1 (3.8) | 4 (12.9) | 2 (5.3) | 0 (0.0) | 2 (66.7) | 4 (26.7) | 1 (2.9) |
| 4-Year Death | 2 (1.2) | 0 (0.0) | 0 (0.0) | 0 (0.0) | 2 (5.3) | 0 (0.0) | 0 (0.0) | 0 (0.0) | 0 (0.0) |
| 5-Year Death  >5-Year Death | 4 (2.4)  8 (4.9) | 1 (7.1)  1 (7.1) | 0 (0.0)  1 (3.8) | 1 (3.2)  2 (6.5) | 1 (2.6)  2 (5.3) | 0 (0.0)  0 (0.0) | 0 (0.0)  0 (0.0) | 1 (6.7)  0 (0.0) | 0 (0.0)  2 (5.7) |
| ^1^n (%) | | | | | | | | | |

**Supplementary** **Table 5: Year wise mortality in Group-B first-line doublet chemo class users as first-line.**

| **Year wise Death** | **Overall**  N = 35^1^ | **Alkylating, Pyrimidines**  N = 5^1^ | **Alkylating, Taxane**  N = 2^1^ | **Antimetabolite, Alkylating**  N = 11^1^ | **Antimetabolite, Tyrosine kinase inhibitor**  N = 3^1^ | **Other Combinations**  N = 9^1^ | **Topoisomerase II Inhibitor, Alkylating**  N = 5^1^ |
| --- | --- | --- | --- | --- | --- | --- | --- |
| Group-C OS |  |  |  |  |  |  |  |
| 1-Year Death | 5 (14.3) | 2 (40.0) | 0 (0.0) | 3 (27.3) | 0 (0.0) | 0 (0.0) | 0 (0.0) |
| 2-Year Death | 13 (37.1) | 2 (40.0) | 1 (50.0) | 5 (45.5) | 0 (0.0) | 3 (33.3) | 2 (40.0) |
| 3-Year Death | 6 (17.1) | 0 (0.0) | 0 (0.0) | 1 (9.1) | 0 (0.0) | 3 (33.3) | 2 (40.0) |
| 4-Year Death | 2 (5.7) | 1 (20.0) | 0 (0.0) | 1 (9.1) | 0 (0.0) | 0 (0.0) | 0 (0.0) |
| 5-Year Death  >5-Year Death | 2 (5.7)  7 (20.0) | 0 (0.0)  0 (0.0) | 1 (50.0)  0 (0.0) | 0 (0.0)  1 (9.1) | 1 (33.3)  2 (66.7) | 0 (0.0)  3 (33.3) | 0 (0.0)  1 (20.0) |
| ^1^n (%) | | | | | | | |

**Supplementary** **Table 6: Year wise mortality in Group-C first-line Doublet chemo class users as first-line.**

**Table7A**

| **Characteristic** | **Overall**  N = 226^1^ | **Alkylating, Anti-mitotic**  N = 6^1^ | **Alkylating, Pyrimidines**  N = 8^1^ | **Alkylating, Taxane**  N = 92^1^ | **Antimetabolite, Alkylating**  N = 61^1^ | **Antimetabolite, Taxane**  N = 1^1^ | **Other Doublet Combination**  N = 42^1^ | **Topoisomerase II Inhibitor, Alkylating**  N = 2^1^ | **Tyrosine kinase inhibitor, Antimetabolite**  N = 14^1^ |
| --- | --- | --- | --- | --- | --- | --- | --- | --- | --- |
| Group A OS |  |  |  |  |  |  |  |  |  |
| 1-Year Death | 172 (76.1) | 2 (33.3) | 7 (87.5) | 65 (70.7) | 53 (86.9) | 1 (100.0) | 36 (85.7) | 2 (100.0) | 6 (42.9) |
| 2-Year Death | 34 (15.0) | 1 (16.7) | 0 (0.0) | 18 (19.6) | 5 (8.2) | 0 (0.0) | 4 (9.5) | 0 (0.0) | 6 (42.9) |
| 3-Year Death | 10 (4.4) | 2 (33.3) | 0 (0.0) | 5 (5.4) | 1 (1.6) | 0 (0.0) | 0 (0.0) | 0 (0.0) | 2 (14.3) |
| 4-Year Death  >5-Year Death | 5 (2.2)  5 (2.2) | 0 (0.0)  1 (16.7) | 1 (12.5)  0 (0.0) | 1 (1.1)  3 (3.3) | 2 (3.3)  0 (0.0) | 0 (0.0)  0 (0.0) | 1 (2.4)  1 (2.4) | 0 (0.0)  0 (0.0) | 0 (0.0)  0 (0.0) |
| ^1^n (%) | | | | | | | | | |

**Table7B**

| **Characteristic** | **Overall**  N = 226^1^ | **Other triplets**  N = 83^1^ | **Triplet Biologics**  N = 49^1^ | **Triplet Pyrimidines**  N = 10^1^ | **Triplet RTKI**  N = 57^1^ | **Triplet Taxane**  N = 22^1^ | **Triplet Topoisomerase inhibitor**  N = 5^1^ |
| --- | --- | --- | --- | --- | --- | --- | --- |
| Group A OS |  |  |  |  |  |  |  |
| 1-Year Death | 172 (76.1) | 61 (73.5) | 41 (83.7) | 6 (60.0) | 40 (70.2) | 20 (90.9) | 4 (80.0) |
| 2-Year Death | 34 (15.0) | 14 (16.9) | 5 (10.2) | 4 (40.0) | 10 (17.5) | 1 (4.5) | 0 (0.0) |
| 3-Year Death | 10 (4.4) | 5 (6.0) | 1 (2.0) | 0 (0.0) | 2 (3.5) | 1 (4.5) | 1 (20.0) |
| 4-Year Death  >5-Year Death | 5 (2.2)  5 (2.2) | 3 (3.6)  0 (0.0) | 0 (0.0)  2 (4.1) | 0 (0.0)  0 (0.0) | 2 (3.5)  3 (5.3) | 0 (0.0)  0 (0.0) | 0 (0.0)  0 (0.0) |
| ^1^n (%) | | | | | | | |

**Supplementary** **Table 7: Year wise mortality in Group-A First-Line Doublet combination of triplet users (Table7A) and overall triplet chemo class type (Table7B). No deaths observed at 5-Year. Biologics, Taxane, Topoisomerase Inhibitor Pyrimidines, and RTKI were continued as triplet combination after end of doublets as first-line chemo (Table7A). Other triplet combinations are prescribed as second line chemotherapy because it consisted of alkylating agents and others types.**

**Table 8A**

| **Year wise Death** | **Overall**  N = 258^1^ | **Alkylating, Anti-mitotic**  N = 29^1^ | **Alkylating, Pyrimidines**  N = 22^1^ | **Alkylating, Taxane**  N = 90^1^ | **Antimetabolite, Alkylating**  N = 62^1^ | **Antimetabolite, Taxane**  N = 1^1^ | **Other Doublet Combination**  N = 29^1^ | **Topoisomerase II Inhibitor, Alkylating**  N = 12^1^ | **Tyrosine kinase inhibitor, Antimetabolite**  N = 13^1^ |
| --- | --- | --- | --- | --- | --- | --- | --- | --- | --- |
| Group B OS |  |  |  |  |  |  |  |  |  |
| 1-Year Death | 114 (44.2) | 4 (13.8) | 19 (86.4) | 37 (41.1) | 34 (54.8) | 0 (0.0) | 7 (24.1) | 10 (83.3) | 3 (23.1) |
| 2-Year Death | 78 (30.2) | 9 (31.0) | 3 (13.6) | 30 (33.3) | 14 (22.6) | 1 (100.0) | 13 (44.8) | 2 (16.7) | 6 (46.2) |
| 3-Year Death | 39 (15.1) | 9 (31.0) | 0 (0.0) | 14 (15.6) | 10 (16.1) | 0 (0.0) | 3 (10.3) | 0 (0.0) | 3 (23.1) |
| 4-Year Death | 20 (7.8) | 5 (17.2) | 0 (0.0) | 6 (6.7) | 3 (4.8) | 0 (0.0) | 6 (20.7) | 0 (0.0) | 0 (0.0) |
| 5-Year Death  >5-Year Death | 5 (1.9)  2 (0.8) | 1 (3.4)  1 (3.4) | 0 (0.0)  0 (0.0) | 2 (2.2)  1 (1.1) | 1 (1.6)  0 (0.0) | 0 (0.0)  0 (0.0) | 0 (0.0)  0 (0.0) | 0 (0.0)  0 (0.0) | 1 (7.7)  0 (0.0) |
| ^1^n (%) | | | | | | | | | |

**Table 8B**

| **Year wise Death** | **Overall**  N = 258^1^ | **Other triplets**  N = 83^1^ | **Triplet Biologics**  N = 56^1^ | **Triplet Pyrimidines**  N = 36^1^ | **Triplet RTKI**  N = 44^1^ | **Triplet Taxane**  N = 29^1^ | **Triplet Topoisomerase inhibitor**  N = 10^1^ |
| --- | --- | --- | --- | --- | --- | --- | --- |
| Group B OS |  |  |  |  |  |  |  |
| 1-Year Death | 114 (44.2) | 21 (25.3) | 34 (60.7) | 21 (58.3) | 12 (27.3) | 19 (65.5) | 7 (70.0) |
| 2-Year Death | 78 (30.2) | 32 (38.6) | 16 (28.6) | 10 (27.8) | 11 (25.0) | 6 (20.7) | 3 (30.0) |
| 3-Year Death | 39 (15.1) | 17 (20.5) | 3 (5.4) | 3 (8.3) | 13 (29.5) | 3 (10.3) | 0 (0.0) |
| 4-Year Death | 20 (7.8) | 11 (13.3) | 0 (0.0) | 1 (2.8) | 7 (15.9) | 1 (3.4) | 0 (0.0) |
| 5-Year Death  >5-Year Death | 5 (1.9)  2 (0.8) | 1 (1.2)  1 (1.2) | 2 (3.6)  1 (1.8) | 1 (2.8)  0 (0.0) | 1 (2.3)  0 (0.0) | 0 (0.0)  0 (0.0) | 0 (0.0)  0 (0.0) |
| ^1^n (%) | | | | | | | |

**Supplementary** **Table 8: Year wise mortality in Group-C First-Line Doublet chemo class of triplet users (Table8A) and overall triplet chemo class type (Table8B). No deaths observed at 5-Year. Biologics, Taxane, Topoisomerase Inhibitor Pyrimidines, and RTKI were continued as triplet combination after end of doublets as first-line chemo (Table8A). Other triplet combinations are prescribed as second line chemotherapy because it consisted of alkylating agents and others types.**

**Table 9A**

| **Year wise Death** | **Overall**  N = 139^1^ | **Alkylating, Anti-mitotic**  N = 11^1^ | **Alkylating, Pyrimidines**  N = 12^1^ | **Alkylating, Taxane**  N = 33^1^ | **Antimetabolite, Alkylating**  N = 20^1^ | **Antimetabolite, Taxane**  N = 3^1^ | **Other Doublet Combination**  N = 30^1^ | **Topoisomerase II Inhibitor, Alkylating**  N = 20^1^ | **Tyrosine kinase inhibitor, Antimetabolite**  N = 10^1^ |
| --- | --- | --- | --- | --- | --- | --- | --- | --- | --- |
| Group C OS |  |  |  |  |  |  |  |  |  |
| 1-Year Death | 18 (12.9) | 0 (0.0) | 4 (33.3) | 4 (12.1) | 1 (5.0) | 0 (0.0) | 4 (13.3) | 5 (25.0) | 0 (0.0) |
| 2-Year Death | 36 (25.9) | 3 (27.3) | 5 (41.7) | 8 (24.2) | 6 (30.0) | 1 (33.3) | 7 (23.3) | 6 (30.0) | 0 (0.0) |
| 3-Year Death | 34 (24.5) | 3 (27.3) | 1 (8.3) | 9 (27.3) | 6 (30.0) | 1 (33.3) | 6 (20.0) | 5 (25.0) | 3 (30.0) |
| 4-Year Death | 25 (18.0) | 1 (9.1) | 2 (16.7) | 7 (21.2) | 2 (10.0) | 0 (0.0) | 7 (23.3) | 3 (15.0) | 3 (30.0) |
| 5-Year Death  >5-Year Death | 4 (2.9)  22 (15.8) | 0 (0.0)  4 (36.4) | 0 (0.0)  0 (0.0) | 1 (3.0)  4 (12.1) | 0 (0.0)  5 (25.0) | 0 (0.0)  1 (33.3) | 1 (3.3)  5 (16.7) | 0 (0.0)  1 (5.0) | 2 (20.0)  2 (20.0) |
| ^1^n (%) | | | | | | | | | |

**Table 9B**

| **Characteristic** | **Overall**  N = 139^1^ | **Other triplets**  N = 42^1^ | **Triplet Biologics**  N = 20^1^ | **Triplet Pyrimidines**  N = 26^1^ | **Triplet RTKI**  N = 20^1^ | **Triplet Taxane**  N = 13^1^ | **Triplet Topoisomerase inhibitor**  N = 18^1^ |
| --- | --- | --- | --- | --- | --- | --- | --- |
| Group C OS |  |  |  |  |  |  |  |
| 1-Year Death | 18 (12.9) | 0 (0.0) | 4 (20.0) | 3 (11.5) | 2 (10.0) | 4 (30.8) | 5 (27.8) |
| 2-Year Death | 36 (25.9) | 7 (16.7) | 4 (20.0) | 13 (50.0) | 3 (15.0) | 4 (30.8) | 5 (27.8) |
| 3-Year Death | 34 (24.5) | 14 (33.3) | 3 (15.0) | 6 (23.1) | 5 (25.0) | 2 (15.4) | 4 (22.2) |
| 4-Year Death | 25 (18.0) | 11 (26.2) | 5 (25.0) | 2 (7.7) | 2 (10.0) | 2 (15.4) | 3 (16.7) |
| 5-Year Death  >5-Year Death | 4 (2.9)  22 (15.8) | 1 (2.4)  9 (21.4) | 1 (5.0)  3 (15.0) | 1 (3.8)  1 (3.8) | 1 (5.0)  7 (35.0) | 0 (0.0)  1 (7.7) | 0 (0.0)  1 (5.6) |
| ^1^n (%) | | | | | | | |

**Supplementary** **Table 9: Year wise mortality in Group-C First-Line Doublet chemo class of triplet users (Table9A) and overall triplet chemo class** type **(Table9B). No deaths observed at 5-Year. Biologics, Taxane, Topoisomerase Inhibitor Pyrimidines, and RTKI were continued as triplet combination after end of doublets as first-line chemo (Table9A). Other triplet combinations are prescribed as second line chemotherapy because it consisted of alkylating agents and others types.**

| **Characteristic** | **Overall**  N = 46^1^ | **Alkylating, Anti-mitotic**  N = 1^1^ | **Alkylating, Antimetabolite**  N = 8^1^ | **Alkylating, Taxane**  N = 12^1^ | **Other combnations as first line doublet**  N = 10^1^ | **Pyrimidines, Alkylating**  N = 3^1^ | **Tyrosine kinase inhibitor, Alkylating**  N = 6^1^ | **Tyrosine kinase inhibitor, Antimetabolite**  N = 6^1^ |
| --- | --- | --- | --- | --- | --- | --- | --- | --- |
| Group A OS |  |  |  |  |  |  |  |  |
| 1-Year Death | 32 (69.6) | 1 (100.0) | 7 (87.5) | 10 (83.3) | 7 (70.0) | 2 (66.7) | 4 (66.7) | 1 (16.7) |
| 2-Year Death | 10 (21.7) | 0 (0.0) | 1 (12.5) | 1 (8.3) | 3 (30.0) | 1 (33.3) | 0 (0.0) | 4 (66.7) |
| 3-Year Death | 2 (4.3) | 0 (0.0) | 0 (0.0) | 1 (8.3) | 0 (0.0) | 0 (0.0) | 0 (0.0) | 1 (16.7) |
| 4-Year Death | 1 (2.2) | 0 (0.0) | 0 (0.0) | 0 (0.0) | 0 (0.0) | 0 (0.0) | 1 (16.7) | 0 (0.0) |
| 5-Year Death | 1 (2.2) | 0 (0.0) | 0 (0.0) | 0 (0.0) | 0 (0.0) | 0 (0.0) | 1 (16.7) | 0 (0.0) |
| ^1^n (%) | | | | | | | | |

**Supplementary** **Table 10: Year wise mortality in Group-A quadruplet users. First-line chemo of quadruplet chemo class users.**

| **Characteristic** | **Overall**  N = 46^1^ | **Alkylating, Antimetabolite**  N = 1^1^ | **Alkylating, Biologics**  N = 4^1^ | **Anti-mitotic, Biologics**  N = 1^1^ | **Antimetabolite, Biologics**  N = 5^1^ | **Antimetabolite, Pyrimidines**  N = 5^1^ | **Antimetabolite, Taxane**  N = 2^1^ | **Antimetabolite, Tyrosine kinase inhibitor**  N = 7^1^ | **Other Combination as quadruplet**  N = 3^1^ | **Pyrimidines, Anti-mitotic**  N = 2^1^ | **Pyrimidines, Taxane**  N = 2^1^ | **Taxane, Alkylating**  N = 2^1^ | **Taxane, Anti-mitotic**  N = 1^1^ | **Taxane, Biologics**  N = 4^1^ | **Taxane, Tyrosine kinase inhibitor**  N = 2^1^ | **Tyrosine kinase inhibitor, Biologics**  N = 4^1^ | **Tyrosine kinase inhibitor, Pyrimidines**  N = 1^1^ |
| --- | --- | --- | --- | --- | --- | --- | --- | --- | --- | --- | --- | --- | --- | --- | --- | --- | --- |
| Group A OS |  |  |  |  |  |  |  |  |  |  |  |  |  |  |  |  |  |
| 1-Year Death | 32 (69.6) | 1 (100.0) | 1 (25.0) | 0 (0.0) | 3 (60.0) | 5 (100.0) | 2 (100.0) | 5 (71.4) | 1 (33.3) | 2 (100.0) | 2 (100.0) | 1 (50.0) | 0 (0.0) | 4 (100.0) | 1 (50.0) | 3 (75.0) | 1 (100.0) |
| 2-Year Death | 10 (21.7) | 0 (0.0) | 2 (50.0) | 1 (100.0) | 0 (0.0) | 0 (0.0) | 0 (0.0) | 1 (14.3) | 2 (66.7) | 0 (0.0) | 0 (0.0) | 1 (50.0) | 1 (100.0) | 0 (0.0) | 1 (50.0) | 1 (25.0) | 0 (0.0) |
| 3-Year Death | 2 (4.3) | 0 (0.0) | 1 (25.0) | 0 (0.0) | 0 (0.0) | 0 (0.0) | 0 (0.0) | 1 (14.3) | 0 (0.0) | 0 (0.0) | 0 (0.0) | 0 (0.0) | 0 (0.0) | 0 (0.0) | 0 (0.0) | 0 (0.0) | 0 (0.0) |
| 4-Year Death | 1 (2.2) | 0 (0.0) | 0 (0.0) | 0 (0.0) | 1 (20.0) | 0 (0.0) | 0 (0.0) | 0 (0.0) | 0 (0.0) | 0 (0.0) | 0 (0.0) | 0 (0.0) | 0 (0.0) | 0 (0.0) | 0 (0.0) | 0 (0.0) | 0 (0.0) |
| 5-Year Death | 1 (2.2) | 0 (0.0) | 0 (0.0) | 0 (0.0) | 1 (20.0) | 0 (0.0) | 0 (0.0) | 0 (0.0) | 0 (0.0) | 0 (0.0) | 0 (0.0) | 0 (0.0) | 0 (0.0) | 0 (0.0) | 0 (0.0) | 0 (0.0) | 0 (0.0) |
| ^1^n (%) | | | | | | | | | | | | | | | | | |

**Supplementary** **Table 11: Year wise mortality in Group-A quadruplet users. Second-line chemo of quadruplet chemo class users.**

| **Characteristic** | **Overall**  N = 192^1^ | **Alkylating, Anti-mitotic**  N = 21^1^ | **Alkylating, Antimetabolite**  N = 50^1^ | **Alkylating, Taxane**  N = 70^1^ | **Topoisomerase II Inhibitor, Alkylating**  N = 5^1^ | **Antimetabolite, Taxane**  N = 1^1^ | **Other combnations as first line doublet**  N = 14^1^ | **Pyrimidines, Alkylating**  N = 9^1^ | **Tyrosine kinase inhibitor, Alkylating**  N = 8^1^ | **Tyrosine kinase inhibitor, Antimetabolite**  N = 14^1^ |
| --- | --- | --- | --- | --- | --- | --- | --- | --- | --- | --- |
| Group B OS |  |  |  |  |  |  |  |  |  |  |
| 1-Year Death | 76 (39.6) | 4 (19.0) | 29 (58.0) | 25 (35.7) | 3 (60.0) | 0 (0.0) | 5 (35.7) | 5 (55.6) | 0 (0.0) | 5 (35.7) |
| 2-Year Death | 60 (31.3) | 6 (28.6) | 13 (26.0) | 22 (31.4) | 2 (40.0) | 1 (100.0) | 6 (42.9) | 4 (44.4) | 4 (50.0) | 2 (14.3) |
| 3-Year Death | 28 (14.6) | 2 (9.5) | 5 (10.0) | 12 (17.1) | 0 (0.0) | 0 (0.0) | 3 (21.4) | 0 (0.0) | 1 (12.5) | 5 (35.7) |
| 4-Year Death | 14 (7.3) | 3 (14.3) | 1 (2.0) | 6 (8.6) | 0 (0.0) | 0 (0.0) | 0 (0.0) | 0 (0.0) | 2 (25.0) | 2 (14.3) |
| 5-Year Death  >5-Year Death | 6 (3.1)  8 (4.2) | 2 (9.5)  4 (19.0) | 2 (4.0)  0 (0.0) | 2 (2.9)  3 (4.3) | 0 (0.0)  0 (0.0) | 0 (0.0)  0 (0.0) | 0 (0.0)  0 (0.0) | 0 (0.0)  0 (0.0) | 0 (0.0)  1 (12.5) | 0 (0.0)  0 (0.0) |
| ^1^n (%) | | | | | | | | | | |

**Supplementary** **Table 12: Year wise mortality in Group-B quadruplet users. First-line chemo of quadruplet chemo class users.**

| **Characteristic** | **Overall**  N = 192^1^ | **Alkylating, Antimetabolite**  N = 4^1^ | **Alkylating, Biologics**  N = 10^1^ | **Alkylating, Pyrimidines**  N = 3^1^ | **Anti-mitotic, Biologics**  N = 17^1^ | **Antimetabolite, Biologics**  N = 11^1^ | **Antimetabolite, Pyrimidines**  N = 14^1^ | **Antimetabolite, Taxane**  N = 7^1^ | **Antimetabolite, Tyrosine kinase inhibitor**  N = 34^1^ | **Biologics, Pyrimidines**  N = 19^1^ | **Other Combination as quadruplet**  N = 12^1^ | **Pyrimidines, Anti-mitotic**  N = 7^1^ | **Pyrimidines, Taxane**  N = 10^1^ | **Taxane, Alkylating**  N = 3^1^ | **Taxane, Anti-mitotic**  N = 2^1^ | **Taxane, Biologics**  N = 19^1^ | **Taxane, Tyrosine kinase inhibitor**  N = 4^1^ | **Tyrosine kinase inhibitor, Biologics**  N = 8^1^ | **Tyrosine kinase inhibitor, Pyrimidines**  N = 8^1^ |
| --- | --- | --- | --- | --- | --- | --- | --- | --- | --- | --- | --- | --- | --- | --- | --- | --- | --- | --- | --- |
| Group B OS |  |  |  |  |  |  |  |  |  |  |  |  |  |  |  |  |  |  |  |
| 1-Year Death | 76 (39.6) | 0 (0.0) | 4 (40.0) | 2 (66.7) | 13 (76.5) | 1 (9.1) | 4 (28.6) | 3 (42.9) | 5 (14.7) | 12 (63.2) | 5 (41.7) | 4 (57.1) | 4 (40.0) | 1 (33.3) | 0 (0.0) | 10 (52.6) | 4 (100.0) | 2 (25.0) | 2 (25.0) |
| 2-Year Death | 60 (31.3) | 3 (75.0) | 1 (10.0) | 0 (0.0) | 1 (5.9) | 4 (36.4) | 5 (35.7) | 2 (28.6) | 12 (35.3) | 4 (21.1) | 5 (41.7) | 3 (42.9) | 4 (40.0) | 2 (66.7) | 2 (100.0) | 5 (26.3) | 0 (0.0) | 4 (50.0) | 3 (37.5) |
| 3-Year Death | 28 (14.6) | 1 (25.0) | 3 (30.0) | 1 (33.3) | 1 (5.9) | 1 (9.1) | 3 (21.4) | 1 (14.3) | 9 (26.5) | 1 (5.3) | 0 (0.0) | 0 (0.0) | 2 (20.0) | 0 (0.0) | 0 (0.0) | 2 (10.5) | 0 (0.0) | 1 (12.5) | 2 (25.0) |
| 4-Year Death | 14 (7.3) | 0 (0.0) | 2 (20.0) | 0 (0.0) | 2 (11.8) | 3 (27.3) | 1 (7.1) | 1 (14.3) | 2 (5.9) | 1 (5.3) | 0 (0.0) | 0 (0.0) | 0 (0.0) | 0 (0.0) | 0 (0.0) | 1 (5.3) | 0 (0.0) | 0 (0.0) | 1 (12.5) |
| 5-Year Death  >5-Year Death | 6 (3.1)  8 (4.2) | 0 (0.0)  0 (0.0) | 0 (0.0)  0 (0.0) | 0 (0.0)  0 (0.0) | 0 (0.0)  0 (0.0) | 0 (0.0)  2 (18.2) | 1 (7.1)  0 (0.0) | 0 (0.0)  0 (0.0) | 2 (5.9)  4 (11.8) | 1 (5.3)  0 (0.0) | 1 (8.3)  1 (8.3) | 0 (0.0)  0 (0.0) | 0 (0.0)  0 (0.0) | 0 (0.0)  0 (0.0) | 0 (0.0)  0 (0.0) | 0 (0.0)  1 (5.3) | 0 (0.0)  0 (0.0) | 1 (12.5)  0 (0.0) | 0 (0.0)  0 (0.0) |
| ^1^n (%) | | | | | | | | | | | | | | | | | | | |

**Supplementary** **Table 13: Year wise mortality in Group-B quadruplet users. Second-line chemo of quadruplet chemo class users.**

| **Characteristic** | **Overall**  N = 331^1^ | **Alkylating, Anti-mitotic**  N = 39^1^ | **Alkylating, Antimetabolite**  N = 68^1^ | **Alkylating, Taxane**  N = 85^1^ | **Alkylating, Topoisomerase II Inhibitor**  N = 8^1^ | **Antimetabolite, Taxane**  N = 8^1^ | **Other combnations as first line doublet**  N = 37^1^ | **Pyrimidines, Alkylating**  N = 22^1^ | **Tyrosine kinase inhibitor, Alkylating**  N = 35^1^ | **Tyrosine kinase inhibitor, Antimetabolite**  N = 29^1^ |
| --- | --- | --- | --- | --- | --- | --- | --- | --- | --- | --- |
| Group C OS |  |  |  |  |  |  |  |  |  |  |
| 1-Year Death | 24 (7.3) | 0 (0.0) | 7 (10.3) | 8 (9.4) | 1 (12.5) | 2 (25.0) | 2 (5.4) | 2 (9.1) | 1 (2.9) | 1 (3.4) |
| 2-Year Death | 91 (27.5) | 4 (10.3) | 25 (36.8) | 18 (21.2) | 5 (62.5) | 3 (37.5) | 11 (29.7) | 10 (45.5) | 8 (22.9) | 7 (24.1) |
| 3-Year Death | 84 (25.4) | 10 (25.6) | 13 (19.1) | 24 (28.2) | 1 (12.5) | 2 (25.0) | 13 (35.1) | 5 (22.7) | 7 (20.0) | 9 (31.0) |
| 4-Year Death | 59 (17.8) | 12 (30.8) | 13 (19.1) | 13 (15.3) | 1 (12.5) | 0 (0.0) | 6 (16.2) | 2 (9.1) | 6 (17.1) | 6 (20.7) |
| 5-Year Death  >5-Year Death | 29 (8.8)  44 (13.3) | 4 (10.3)  9 (23.1) | 2 (2.9)  8 (11.8) | 6 (7.1)  16 (18.8) | 0 (0.0)  0 (0.0) | 1 (12.5)  0 (0.0) | 1 (2.7)  4 (10.8) | 2 (9.1)  1 (4.5) | 8 (22.9)  5 (14.3) | 5 (17.2)  1 (3.4) |
| ^1^n (%) | | | | | | | | | | |

**Supplementary** **Table 14: Year wise mortality in Group-C quadruplet users. First-line chemo of quadruplet chemo class users.**

| **Characteristic** | | **Overall**  N = 331^1^ | **Alkylating, Antimetabolite**  N = 6^1^ | **Alkylating, Biologics**  N = 19^1^ | **Alkylating, Pyrimidines**  N = 15^1^ | **Anti-mitotic, Biologics**  N = 15^1^ | **Antimetabolite, Biologics**  N = 28^1^ | **Antimetabolite, Pyrimidines**  N = 19^1^ | **Antimetabolite, Taxane**  N = 9^1^ | **Antimetabolite, Tyrosine kinase inhibitor**  N = 41^1^ | **Biologics, Pyrimidines**  N = 35^1^ | **Other Combination as quadruplet**  N = 40^1^ | **Other Combination as second line quadruplet**  N = 1^1^ | **Pyrimidines, Anti-mitotic**  N = 15^1^ | **Pyrimidines, Taxane**  N = 14^1^ | **Taxane, Alkylating**  N = 11^1^ | **Taxane, Anti-mitotic**  N = 10^1^ | **Taxane, Biologics**  N = 20^1^ | **Taxane, Tyrosine kinase inhibitor**  N = 14^1^ | **Tyrosine kinase inhibitor, Biologics**  N = 6^1^ | **Tyrosine kinase inhibitor, Pyrimidines**  N = 13^1^ |
| --- | --- | --- | --- | --- | --- | --- | --- | --- | --- | --- | --- | --- | --- | --- | --- | --- | --- | --- | --- | --- | --- |
| Group C OS | |  |  |  |  |  |  |  |  |  |  |  |  |  |  |  |  |  |  |  |  |
| 1-Year Death | | 24 (7.3) | 0 (0.0) | 0 (0.0) | 2 (13.3) | 1 (6.7) | 1 (3.6) | 3 (15.8) | 0 (0.0) | 2 (4.9) | 4 (11.4) | 3 (7.5) | 0 (0.0) | 3 (20.0) | 1 (7.1) | 0 (0.0) | 1 (10.0) | 1 (5.0) | 1 (7.1) | 0 (0.0) | 1 (7.7) |
| 2-Year Death | | 91 (27.5) | 1 (16.7) | 5 (26.3) | 6 (40.0) | 7 (46.7) | 3 (10.7) | 6 (31.6) | 4 (44.4) | 6 (14.6) | 5 (14.3) | 13 (32.5) | 0 (0.0) | 5 (33.3) | 4 (28.6) | 4 (36.4) | 4 (40.0) | 8 (40.0) | 7 (50.0) | 1 (16.7) | 2 (15.4) |
| 3-Year Death | | 84 (25.4) | 2 (33.3) | 5 (26.3) | 5 (33.3) | 1 (6.7) | 7 (25.0) | 4 (21.1) | 2 (22.2) | 10 (24.4) | 11 (31.4) | 8 (20.0) | 1 (100.0) | 5 (33.3) | 4 (28.6) | 3 (27.3) | 3 (30.0) | 4 (20.0) | 3 (21.4) | 0 (0.0) | 6 (46.2) |
| 4-Year Death | | 59 (17.8) | 0 (0.0) | 5 (26.3) | 1 (6.7) | 3 (20.0) | 5 (17.9) | 2 (10.5) | 1 (11.1) | 8 (19.5) | 10 (28.6) | 8 (20.0) | 0 (0.0) | 0 (0.0) | 4 (28.6) | 3 (27.3) | 1 (10.0) | 5 (25.0) | 1 (7.1) | 2 (33.3) | 0 (0.0) |
| 5-Year Death  >5-Year Death | | 29 (8.8)  44 (13.3) | 1 (16.7)  2 (33.3) | 2 (10.5)  2 (10.5) | 0 (0.0)  1 (6.7) | 0 (0.0)  3 (20.0) | 7 (25.0)  5 (17.9) | 2 (10.5)  2 (10.5) | 2 (22.2)  0 (0.0) | 4 (9.8)  11 (26.8) | 0 (0.0)  5 (14.3) | 5 (12.5)  3 (7.5) | 0 (0.0)  0 (0.0) | 1 (6.7)  1 (6.7) | 0 (0.0)  1 (7.1) | 1 (9.1)  0 (0.0) | 0 (0.0)  1 (10.0) | 1 (5.0)  1 (5.0) | 1 (7.1)  1 (7.1) | 2 (33.3)  1 (16.7) | 0 (0.0)  4 (30.8) |
| ^1^n (%) |  |  |  |  |  |  |  |  |  |  |  |  |  |  |  |  |  |  |  |  |  |

**Supplementary** **Table 15: Year wise mortality in Group-C quadruplet users. Second-line chemo of quadruplet chemo class users.**

| **Year Wise Deaths** | **Comorbidity Yes** | **Comorbidity No** | **No diabetes** | **Pre cancer diabetes** | **Post cancer diabetes** | **No anemia** | **Pre cancer anemia** | **Post cancer anemia** | **No Anxiety** | **Pre cancer anxiety** | **Post cancer anxiety** | **No CAD** | **Pre cancer CAD** | **Post cancer CAD** | **No CLD** | **Pre cancer CLD** | **Post cancer CLD** | **No COPD** | **Pre cancer COPD** | **Post cancer COPD** |
| --- | --- | --- | --- | --- | --- | --- | --- | --- | --- | --- | --- | --- | --- | --- | --- | --- | --- | --- | --- | --- |
| Group-A | N = 542^1^ | N = 747^1^ | N = 1,107^1^ | N = 55^1^ | N = 127^1^ | N = 1,220^1^ | N = 15^1^ | N = 54^1^ | N = 1,202^1^ | N = 17^1^ | N = 70^1^ | N = 1,209^1^ | N = 54^1^ | N = 26^1^ | N = 1,269^1^ | N = 1^1^ | N = 19^1^ | N = 1,084^1^ | N = 67^1^ | N = 138^1^ |
| 1-Year Deaths | 421 (77.7) | 581 (77.8) | 861 (77.8) | 44 (80.0) | 97 (76.4) | 950 (77.9) | 10 (66.7) | 42 (77.8) | 938 (78.0) | 15 (88.2) | 49 (70.0) | 934 (77.3) | 45 (83.3) | 23 (88.5) | 987 (77.8) | 1 (100.0) | 14 (73.7) | 844 (77.9) | 61 (91.0) | 97 (70.3) |
| 2-Year Deaths | 80 (14.8) | 110 (14.7) | 165 (14.9) | 6 (10.9) | 19 (15.0) | 178 (14.6) | 4 (26.7) | 8 (14.8) | 174 (14.5) | 2 (11.8) | 14 (20.0) | 184 (15.2) | 5 (9.3) | 1 (3.8) | 187 (14.7) | 0 (0.0) | 3 (15.8) | 156 (14.4) | 3 (4.5) | 31 (22.5) |
| 3-Year Deaths | 21 (3.9) | 30 (4.0) | 43 (3.9) | 3 (5.5) | 5 (3.9) | 47 (3.9) | 1 (6.7) | 3 (5.6) | 47 (3.9) | 0 (0.0) | 4 (5.7) | 48 (4.0) | 2 (3.7) | 1 (3.8) | 50 (3.9) | 0 (0.0) | 1 (5.3) | 44 (4.1) | 1 (1.5) | 6 (4.3) |
| 4-Year Deaths | 9 (1.7) | 15 (2.0) | 21 (1.9) | 0 (0.0) | 3 (2.4) | 24 (2.0) | 0 (0.0) | 0 (0.0) | 22 (1.8) | 0 (0.0) | 2 (2.9) | 24 (2.0) | 0 (0.0) | 0 (0.0) | 23 (1.8) | 0 (0.0) | 1 (5.3) | 23 (2.1) | 1 (1.5) | 0 (0.0) |
| 5-Year Deaths  >6-Year Deaths  Group-B  1-Year Deaths  2-Year Deaths  3-Year Deaths  4-Year Deaths  5-Year Deaths  >6-Year Deaths  Group-C  1-Year Deaths  2-Year Deaths  3-Year Deaths  4-Year Deaths  5-Year Deaths  >6-Year Deaths | 2 (0.4)  9 (1.7)  N = 338^1^  127 (41.0)  91 (29.4)  46 (14.8)  25 (8.1)  9 (2.9)  12 (3.9)  N = 284^1^  21 (7.4)  68 (23.9)  63 (22.2)  54 (19.0)  20 (7.0)  58 (20.4) | 5 (0.7)  6 (0.8)  N = 310^1^  167 (49.4)  92 (27.2)  43 (12.7)  19 (5.6)  9 (2.7)  8 (2.4)  N = 238^1^  27 (11.3)  74 (31.1)  63 (26.5)  33 (13.9)  16 (6.7)  25 (10.5) | 6 (0.5)  11 (1.0)  N = 535^1^  248 (46.4)  148 (27.7)  75 (14.0)  33 (6.2)  16 (3.0)  15 (2.8)  N = 421^1^  40 (9.5)  121 (28.7)  107 (25.4)  63 (15.0)  30 (7.1)  60 (14.3) | 0 (0.0)  2 (3.6)  N = 30^1^  14 (46.7)  7 (23.3)  5 (16.7)  3 (10.0)  0 (0.0)  1 (3.3)  N = 24^1^  3 (12.5)  6 (25.0)  5 (20.8)  6 (25.0)  0 (0.0)  4 (16.7) | 1 (0.8)  2 (1.6)  N = 83^1^  32 (38.6)  28 (33.7)  9 (10.8)  8 (9.6)  2 (2.4)  4 (4.8)  N = 77^1^  5 (6.5)  15 (19.5)  14 (18.2)  18 (23.4)  6 (7.8)  19 (24.7) | 7 (0.6)  14 (1.1)  N = 608^1^  274 (45.1)  174 (28.6)  79 (13.0)  43 (7.1)  18 (3.0)  20 (3.3)  N = 465^1^  43 (9.2)  127 (27.3)  112 (24.1)  76 (16.3)  35 (7.5)  72 (15.5) | 0 (0.0)  0 (0.0)  N = 2^1^  1 (50.0)  1 (50.0)  0 (0.0)  0 (0.0)  0 (0.0)  0 (0.0)  N = 3^1^  1 (33.3)  2 (66.7)  0 (0.0)  0 (0.0)  0 (0.0)  0 (0.0) | 0 (0.0)  1 (1.9)  N = 38^1^  19 (50.0)  8 (21.1)  10 (26.3)  1 (2.6)  0 (0.0)  0 (0.0)  N = 54^1^  4 (7.4)  13 (24.1)  14 (25.9)  11 (20.4)  1 (1.9)  11 (20.4) | 6 (0.5)  15 (1.2)  N = 590^1^  273 (46.3)  165 (28.0)  80 (13.6)  38 (6.4)  16 (2.7)  18 (3.1)  N = 474^1^  45 (9.5)  129 (27.2)  118 (24.9)  79 (16.7)  33 (7.0)  70 (14.8) | 0 (0.0)  0 (0.0)  N = 6^1^  3 (50.0)  2 (33.3)  1 (16.7)  0 (0.0)  0 (0.0)  0 (0.0)  N = 6^1^  0 (0.0)  1 (16.7)  1 (16.7)  2 (33.3)  0 (0.0)  2 (33.3) | 1 (1.4)  0 (0.0)  N = 52^1^  18 (34.6)  16 (30.8)  8 (15.4)  6 (11.5)  2 (3.8)  2 (3.8)  N = 42^1^  3 (7.1)  12 (28.6)  7 (16.7)  6 (14.3)  3 (7.1)  11 (26.2) | 7 (0.6)  12 (1.0)  N = 1,206^1^  550 (45.6)  340 (28.2)  170 (14.1)  76 (6.3)  32 (2.7)  38 (3.2)  N = 482^1^  44 (9.1)  132 (27.4)  115 (23.9)  82 (17.0)  31 (6.4)  78 (16.2) | 0 (0.0)  2 (3.7)  N = 54^1^  28 (51.9)  20 (37.0)  2 (3.7)  2 (3.7)  2 (3.7)  0 (0.0)  N = 21^1^  3 (14.3)  6 (28.6)  7 (33.3)  1 (4.8)  3 (14.3)  1 (4.8) | 0 (0.0)  1 (3.8)  N = 36^1^  10 (27.8)  6 (16.7)  6 (16.7)  10 (27.8)  2 (5.6)  2 (5.6)  N = 19^1^  1 (5.3)  4 (21.1)  4 (21.1)  4 (21.1)  2 (10.5)  4 (21.1) | 7 (0.6)  15 (1.2)  N = 1,270^1^  578 (45.5)  360 (28.3)  172 (13.5)  88 (6.9)  34 (2.7)  38 (3.0)  N = 498^1^  46 (9.2)  140 (28.1)  118 (23.7)  82 (16.5)  35 (7.0)  77 (15.5) | 0 (0.0)  0 (0.0)  N = 0^1^  0 (0.0)  0 (0.0)  0 (0.0)  0 (0.0)  0 (0.0)  0 (0.0)  N = 0^1^  0 (0.0)  0 (0.0)  0 (0.0)  0 (0.0)  0 (0.0)  0 (0.0) | 0 (0.0)  0 (0.0)  N = 26^1^  10 (38.5)  6 (23.1)  6 (23.1)  0 (0.0)  2 (7.7)  2 (7.7)  N = 24^1^  2 (8.3)  2 (8.3)  8 (33.3)  5 (20.8)  1 (4.2)  6 (25.0) | 7 (0.6)  10 (0.9)  N = 542^1^  243 (44.8)  154 (28.4)  77 (14.2)  38 (7.0)  16 (3.0)  14 (2.6)  N = 450^1^  44 (9.8)  124 (27.6)  111 (24.7)  70 (15.6)  31 (6.9)  70 (15.6) | 0 (0.0)  1 (1.5)  N = 22^1^  10 (45.5)  8 (36.4)  1 (4.5)  1 (4.5)  1 (4.5)  1 (4.5)  N = 15^1^  0 (0.0)  4 (26.7)  5 (33.3)  4 (26.7)  0 (0.0)  2 (13.3) | 0 (0.0)  4 (2.9)  N = 84^1^  41 (48.8)  21 (25.0)  11 (13.1)  5 (6.0)  1 (1.2)  5 (6.0)  N = 57^1^  4 (7.0)  14 (24.6)  10 (17.5)  13 (22.8)  5 (8.8)  11 (19.3) |

**Supplementary** **Table 16:** Year wise mortality rate in Group-A, B, & C as per comorbidities diagnosed pre/post primary lung cancer. Mortality rate was calculated post-chemotherapy initiation. In this study 80% patients were diagnosed with Stage III/IV primary lung cancer, most of pre-cancer comorbidities might have occurred during initial cancer because 80% of the primary cancer diagnosed in later stages. Comorbidities can induce cancer here it is very rare case; comorbidities can co-occur with initial diagnosis of cancer or could be chemotherapy induced.
